# Functional PD-1/PD-L1 engagement defines a spatial biomarker of immunotherapy response

**DOI:** 10.64898/2026.04.15.26350929

**Authors:** Tony Ullman, David Krantz, Christophe Avenel, Maria Lung, Fernanda Costa Svedman, Karin Holmsten, Päivi Östling, Anders Ullén, Charlotte Stadler

## Abstract

Effective predictive biomarkers for immune checkpoint inhibitor (ICI) therapy remain an unmet need across solid tumors. Here, we present an integrated spatial proteomics workflow that combines *in situ* proximity ligation assay with multiplexed immunofluorescence to directly resolve PD-1/PD-L1 signaling events at the level of defined cellular phenotypes and their spatial organization within intact tumor tissue.

Applied as a proof-of-concept to tumor samples from patients with metastatic urothelial carcinoma treated with pembrolizumab, this approach reveals that PD-1/PD-L1 interactions specifically involving cytotoxic CD8⁺ T cells are significantly enriched in complete responders, while such interactions are rare in patients with progressive disease. This interaction-defined T cell subset achieves superior discrimination of clinical response compared to single-marker PD-L1 expression or immune cell abundance alone.

By integrating direct detection of protein–protein interactions with high-dimensional single-cell phenotyping, our workflow provides a mechanistically informed, spatially resolved biomarker of functional immune engagement. Beyond urothelial carcinoma, this platform establishes a generalizable framework for translating spatial signaling biology into predictive tools for immunotherapy response across tumor types.

**One Sentence Summary:** *In situ* detection of PD1/PD-L1 interactions between cancer cells and cytotoxic T cells predicts response to immunotherapy in urothelial carcinoma

## INTRODUCTION

Immune checkpoint inhibitor (ICI) therapy has been a game changer for cancer treatment over the past decades, delivering durable clinical benefit across multiple tumor types (1). The most widely used ICIs block the interaction between programmed cell death receptor 1 (PD-1) and its ligand programmed death-ligand 1 (PD-L1). In several solid malignancies, ICIs targeting the PD-1/PD-L1 axis is approved as a perioperative treatment for localized disease or for metastatic disease. For some therapeutic indications, treatment eligibility is restricted to patients with PD-L1–positive tumors, as determined by drug-specific companion immunohistochemical assays. Although PD-L1 testing has entered routine clinical diagnostics, interpretation remains challenging because several non-interchangeable assays and divergent scoring algorithms are in use (e.g., tumor-cell vs. immune-cell scoring or combined positive score). Consequently, the prognostic and predictive value of PD-L1 expression remains inconsistent across studies(2). For example, in advanced urothelial cancer, fewer than 50 % of patients with PD-L1–positive tumors respond to therapy, with overall response rates (ORR) of 20–40 % (3–5). Better predictive biomarkers are needed to refine patient selection, enabling more accurate identification of those most likely to benefit while minimizing ineffective treatment and toxicity in non-responders.

In addition to technical inconsistencies, the complexity of PD-1/PD-L1 signaling — driven by context-dependent expression and diverse cellular interactions — likely further contributes to the limited and variable performance of PD-L1 as a stand-alone biomarker. This complexity is reflected not only in the regulation of PD-L1 expression but also in the dynamic nature of its interaction with PD-1 across distinct cellular compartments within the tumor microenvironment (6).

Because immune checkpoint inhibitors target PD-1–PD-L1 signaling rather than PD-L1 expression itself, the presence of these interactions may provide stronger predictive value than PD-L1 levels alone.

Several studies have attempted to directly detect PD-1–PD-L1 interactions, mainly in lung cancer (7,8). These analyses suggest that the presence and frequency of PD-1/PD-L1 interactions can predict response to ICI therapy more accurately than PD-L1 expression alone. However, these approaches are limited by their focus on the interaction itself, without considering the cellular context in which it occurs. Information on the identity, abundance, and spatial organization of the interacting cell types is largely missing - factors likely critical for interpreting the biological and clinical significance of PD-1–PD-L1 signaling.

In response to interferon-γ, tumor cells upregulate PD-L1, which binds PD-1 on activated T cells and establishes a negative feedback loop that limits T-cell effector activity (9). In solid malignancies such as urothelial cancer, PD-L1 is expressed on both tumor and immune cells, enabling PD-1–PD-L1 signaling between tumor and immune compartments as well as among immune cells themselves (6). The relative contribution of tumor- versus immune-cell PD-L1 may be especially relevant in this highly immunogenic disease, which is characterized by a high mutational burden and dense immune infiltration (10). These features make urothelial cancer a suitable model for biomarker discovery in the context of immune checkpoint blockade.

We reasoned that such complex tumor–immune dynamics demand biomarkers that are both mechanistically informative and analytically robust. Because certain immune-cell subsets play a dominant role in tumor control, resolving their spatial distribution and functional state is particularly critical. For instance, in urothelial cancer, tumors enriched for intratumoral CD8 ⁺ T-effector signatures have been reported to show greater likelihood of response to PD-L1 blockade with atezolizumab (11). Therefore, combining direct assessment of PD-1–PD-L1 interactions with high resolution immune-cell phenotyping may yield more powerful and biologically meaningful predictive biomarkers.

Over the past decade, multiplexed imaging technologies have substantially improved our understanding of the tumor microenvironment as it allows for simultaneous identification of different cell types, their spatial distribution and cellular neighborhoods in situ (12–15), reflecting the rapid maturation of spatial proteomics as a field (16). Although spatial proximity is often used as a surrogate for cell–cell interaction, physical adjacency alone does not demonstrate functional communication. Demonstrating true cell–cell interactions, such as signaling events, requires a direct analysis of protein–protein interactions.

Here, we have developed an integrated assay combining the *in situ* Proximity Ligation Assay (isPLA) technology for detection of PD-1/PD-L1 interactions, with multiplexed immunofluorescence (mIF). This approach enables simultaneous mapping of receptor–ligand interactions to specific cellular phenotypes and their spatial organization within the tumor immune microenvironment.

## RESULTS

### An integrated assay for detecting signaling events between individual cells

To explore PD-1/PD-L1 interaction as a biomarker for response to ICI therapy, we developed an integrated assay combining *in situ* Proximity Ligation Assay (isPLA) with multiplexed immunofluorescence on the PhenoCycler platform. The proximity ligation technology uses pairs of antibodies that bind to protein targets expected to interact. The antibodies are tagged with DNA oligos that hybridize upon close proximity to form a circular DNA product which is subsequently amplified and detected using a fluorescently labeled complementary detection oligo. (DO). Fluorescence is detected only when the targets lie within ∼30–40 nm of each other, enabling highly specific and sensitive detection of protein interactions (17,18).

We applied this approach to target PD-1 and PD-L1 in tumor sections from patients with metastatic urothelial carcinoma (mUC). However, to move beyond detecting PD-1/PD-L1 interactions between neighboring cells without knowledge of their identity, we sought to achieve resolution at the cell phenotype level. To this end, we integrated isPLA with a 14-marker multiplex antibody panel that we previously optimized on the PhenoCycler platform **(Fig. 1).** The platform uses DNA-barcoded antibodies with iterative cycles of reporter hybridization, imaging, and removal, enabling sequential readout across multiple channels and thereby highly multiplexed imaging.

**Fig. 1.**
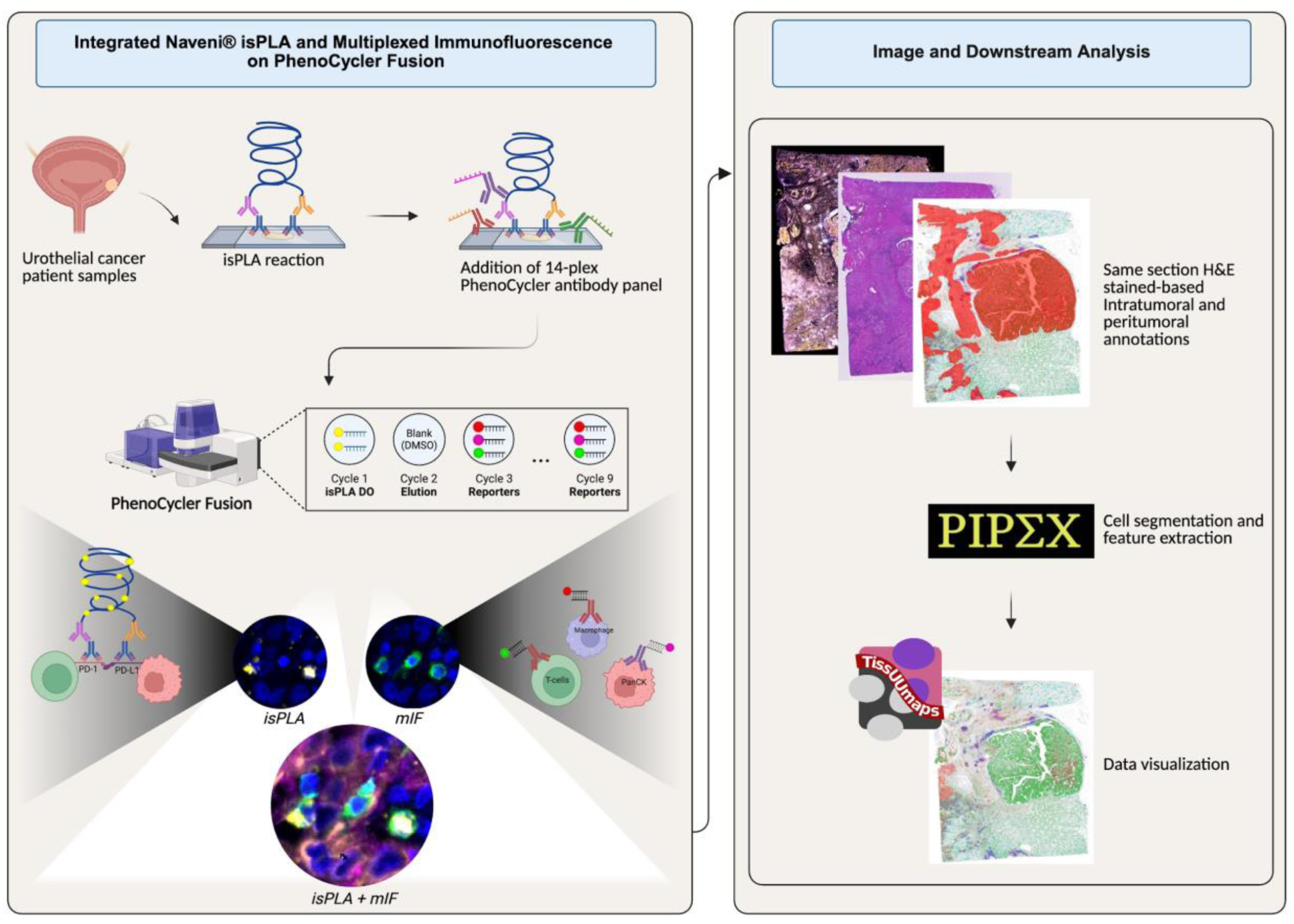
Overview of the integrated isPLA–PhenoCycler workflow. (Left panel) PD-1/PD-L1 in situ PLA was combined with a 14-plex PhenoCycler antibody panel on urothelial cancer tissue sections, followed by iterative staining and imaging. (Right panel) Cells were segmented and tumor and peritumoral regions were annotated based on morphology. The resulting spatially resolved single-cell data were analyzed quantitatively and spatially. Created in BioRender. Ullman, T. (2026) https://BioRender.com/veyfvrd.

To enable detection of the isPLA signal within the multiplexed IF workflow, the signal was detected in the initial PhenoCycler imaging cycle and subsequently removed prior to multiplexed phenotyping. With this setup, one single experimental dataset with integrated data was created, allowing for detection of protein interactions from the isPLA assay, with deep characterization of the tissue microenvironment as detected by the conjugated PhenoCycler antibodies. Further, post-imaging hematoxylin and eosin (H&E) staining was performed to enable morphology-based annotation of tumoral and peritumoral areas. Integrated single-image data were analyzed using the open-source tool PIPEX and visualized using the web-based platform TissUUmaps (17–19). The integrated workflow showed no detectable cross-reactivity between the isPLA detection chemistry and the PhenoCycler antibody panel **(Supp Fig 1).** To assess whether incorporation of the upstream isPLA assay affected antibody performance, paired tissue sections from the same specimen were processed either using a standard PhenoCycler workflow or with the additional isPLA step. Qualitative assessment revealed no differences in tissue integrity between conditions. Quantitative analysis further showed comparable signal-to-noise ratios across all PhenoCycler antibodies, indicating preserved binding performance and signal intensity following isPLA integration.

Positive and negative biological controls confirmed specific isPLA signal detection without evidence of false-positive interactions. **(Supp Fig 1E).**

### Validation of integrated isPLA–mIF workflow in a urothelial cancer tissue section

Following assay integration and validation, we applied the workflow to a urothelial cancer (UC) tissue section enriched for viable tumor and adjacent tertiary lymphoid structures (TLSs) (**Fig. 2**). Selected regions of interest (ROIs) highlight a TLS (ROI 1) and a tumor area (ROI 2). Using isPLA, we detected in situ PD-1/PD-L1 interactions in distinct cellular contexts within the tumor microenvironment. In ROI 1, isPLA signals were observed throughout the TLS, indicating PD-1/PD-L1 engagement within organized immune cell aggregates. In ROI 2, isPLA signals were localized to the tumor–immune interface, consistent with checkpoint interactions involving infiltrating immune cells and tumor cells.

**Fig. 2.**
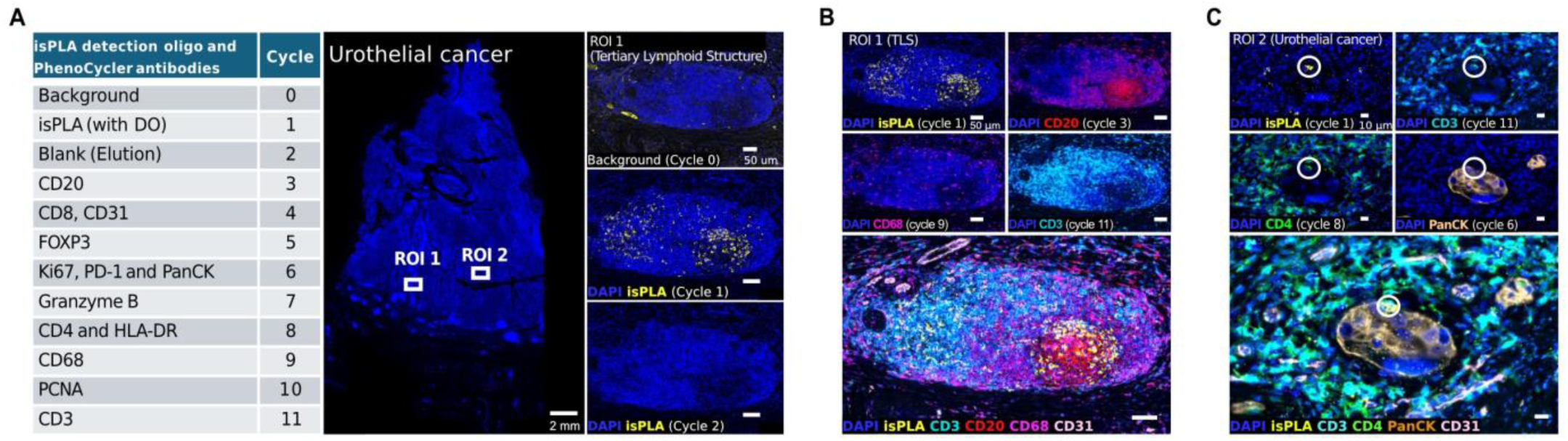
Workflow integrating PD-1/PD-L1 isPLA with multiplexed PhenoCycler antibody detection. (A) Urothelial cancer section was first stained with PD-1/PD-L1 in situ PLA, and the signal was read out by incorporating detection oligos (Atto647) as the first imaging cycle on the PhenoCycler Fusion. Subsequent cycles (summarized in the table) included iterative antibody detection and elution across a 14-plex panel (e.g., CD20, CD31, FOXP3, Ki67, CD68, CD3). The whole-section overview highlights two regions of interest (ROI 1: tertiary lymphoid structure, ROI 2: tumor nest), with representative cycle images showing isPLA signal, blank/elution, and subsequent antibody detections. (B) ROI 1 demonstrates organized lymphoid architecture, where isPLA signal (cycle 1) can be analyzed in the context of immune markers across later cycles (e.g., CD20, CD68, CD3, CD31), visualized in composite overlays. (C) ROI 2 shows isPLA signal localized to CD3⁺ and CD4⁺ T cells interacting with PanCK⁺ tumor cells, alongside CD31⁺ vasculature. This illustrates how the isPLA signal, captured as part of the iterative PhenoCycler workflow, enables combined visualization of protein interactions and immune phenotypes within the same section.

To enable cellular attribution of the isPLA signal, we applied a 14-plex PhenoCycler multiplex immunofluorescence (mIF) panel designed to capture tumor identity, immune cell composition, and functional states. Tumor cells were identified using a pan-cytokeratin (PanCK) antibody cocktail and morphologically verified by a certified uropathologist. CD31 delineated vasculature and stromal architecture. Major immune lineages were defined using CD3, CD4, CD8, CD20, CD68, and FOXP3, while functional markers included PD-L1, HLA-DR, Granzyme B, and Ki-67. mIF analysis of ROI 1 revealed a structured TLS composed of B cells, T cells, macrophages, and associated vasculature. In ROI 2, PanCK⁺ tumor cells were surrounded by a T cell–enriched immune infiltrate.

Integration of isPLA with mIF enabled direct assignment of PD-1/PD-L1 interactions to specific cell types and microanatomical niches. Within the TLS, isPLA signals were predominantly associated with immune cell–immune cell interactions, particularly in B cell–rich regions. In contrast, in the tumor ROI, isPLA signal co-localized with CD3⁺CD4⁺ PD-1–expressing T cells engaging PD-L1–expressing PanCK⁺ tumor cells. Together, these results demonstrate that the integrated isPLA–mIF workflow resolves distinct cellular and spatial contexts of PD-1/PD-L1 engagement in situ, enabling discrimination between checkpoint interactions occurring within immune aggregates and those at the tumor–immune interface.

### PD-1/PD-L1 interactions in pembrolizumab-treated advanced urothelial cancer

To assess the biomarker potential of PD-1/PD-L1 interactions in a spatial, cell-type–resolved context, we analyzed a retrospective cohort of 107 patients with metastatic urothelial cancer (mUC) treated with pembrolizumab. From this cohort, 15 cases were selected (5 complete responses [CR], 10 progressive disease [PD]), balanced for tumor PD-L1 status. Two cases (one CR, one PD) were excluded due to insufficient tumor material, yielding 13 evaluable samples. All specimens were derived from primary tumors except one metastatic sample (**Tables 1–2**).

**Table 1.**
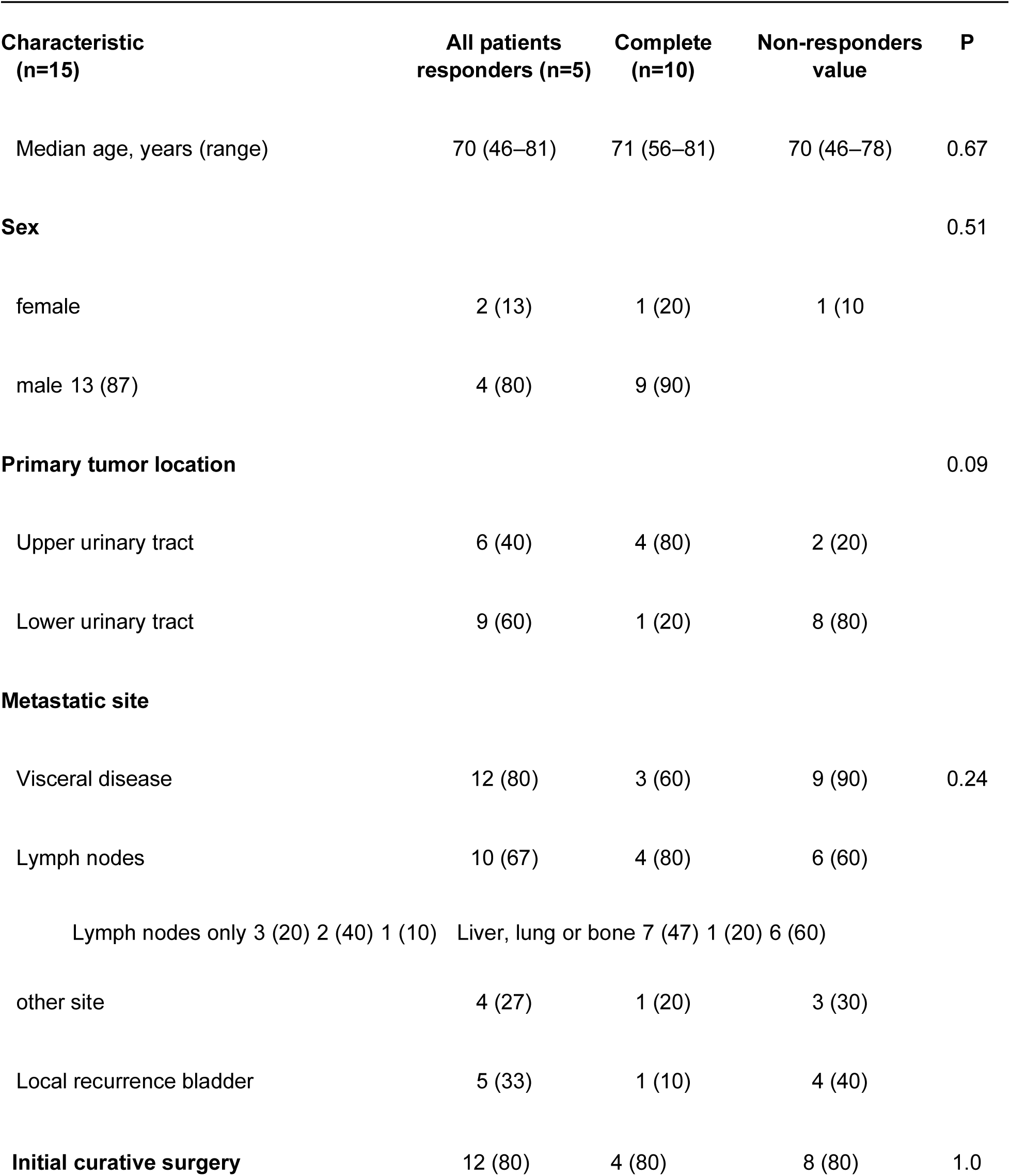
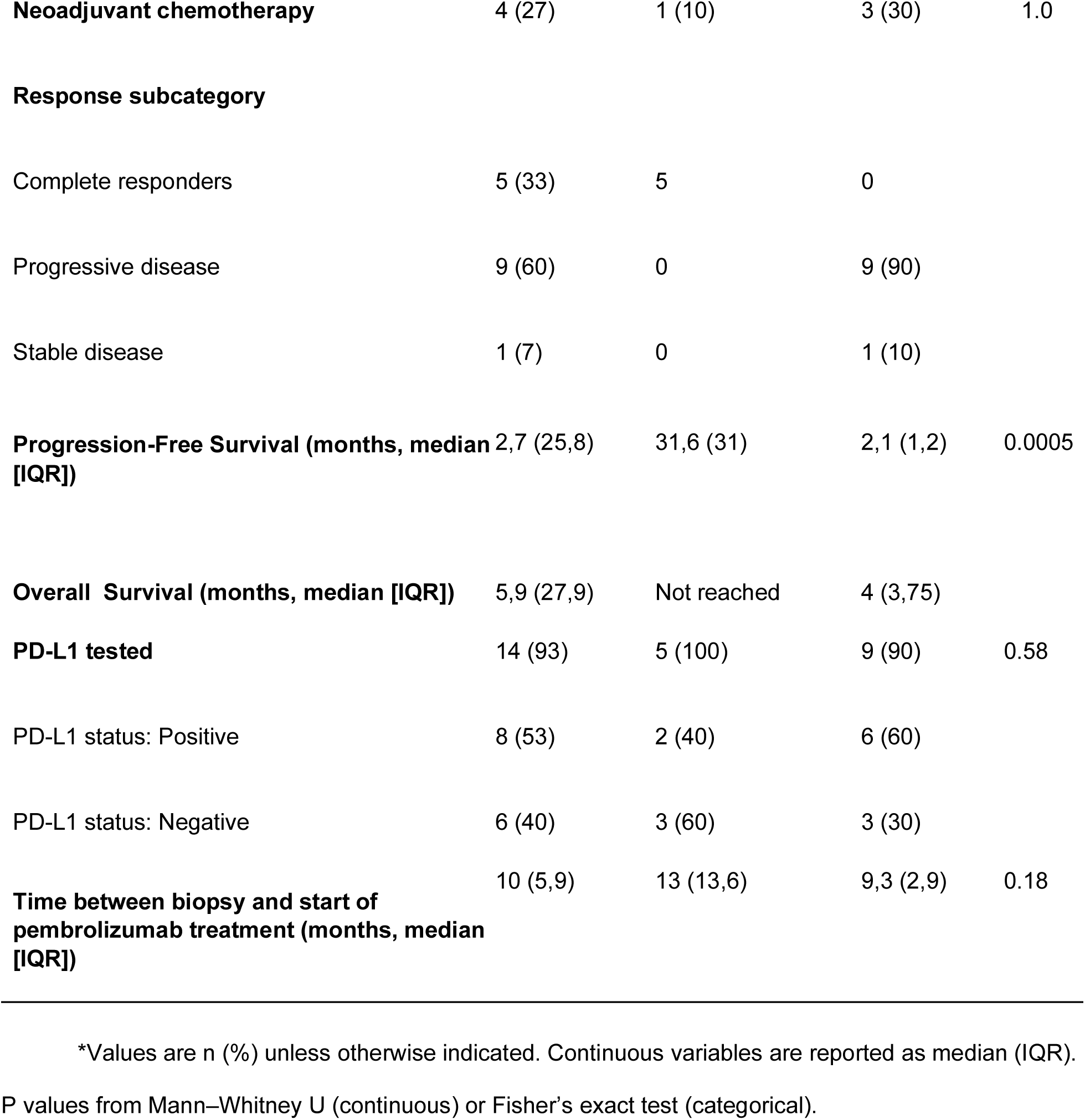
Baseline Characteristics of the Patients.

| 646 |  |  |  |  |  |
| --- | --- | --- | --- | --- | --- |
| 647 | **Characteristic** | **All patients** | **Complete** | **Non-responders** | **P** |
| 648 | **(n=15)** | **responders (n=5)** | **(n=10)** | **value** |  |
| 649 | Median age, years (range) | 70 (46–81) | 71 (56–81) | 70 (46–78) | 0.67 |
| 650 | **Sex** |  |  |  | 0.51 |
| 651 | female | 2 (13) | 1 (20) | 1 (10) |  |
| 652 | male 13 (87) | 4 (80) | 9 (90) |  |  |
| 653 | **Primary tumor location** |  |  |  | 0.09 |
| 654 | Upper urinary tract | 6 (40) | 4 (80) | 2 (20) |  |
| 655 | Lower urinary tract | 9 (60) | 1 (20) | 8 (80) |  |
| 656 | **Metastatic site** |  |  |  |  |
| 657 | Visceral disease | 12 (80) | 3 (60) | 9 (90) | 0.24 |
| 658 | Lymph nodes | 10 (67) | 4 (80) | 6 (60) |  |
| 659 | Lymph nodes only 3 (20) 2 (40) 1 (10) | Liver, lung or bone 7 (47) 1 (20) 6 (60) |  |  |  |
| 660 | other site | 4 (27) | 1 (20) | 3 (30) |  |
| 661 | Local recurrence bladder | 5 (33) | 1 (10) | 4 (40) |  |
|  | **Initial curative surgery** | 12 (80) | 4 (80) | 8 (80) | 1.0 |

|  |  |  |  |  |
| --- | --- | --- | --- | --- |
| **Neoadjuvant chemotherapy** | 4 (27) | 1 (10) | 3 (30) | 1.0 |
| **Response subcategory** |  |  |  |  |
| Complete responders | 5 (33) | 5 | 0 |  |
| Progressive disease | 9 (60) | 0 | 9 (90) |  |
| Stable disease | 1 (7) | 0 | 1 (10) |  |
| **Progression-Free Survival (months, median [IQR])** | 2,7 (25,8) | 31,6 (31) | 2,1 (1,2) | 0.0005 |
| **Overall Survival (months, median [IQR])** | 5,9 (27,9) | Not reached | 4 (3,75) |  |
| **PD-L1 tested** | 14 (93) | 5 (100) | 9 (90) | 0.58 |
| PD-L1 status: Positive | 8 (53) | 2 (40) | 6 (60) |  |
| PD-L1 status: Negative | 6 (40) | 3 (60) | 3 (30) |  |
| **Time between biopsy and start of pembrolizumab treatment (months, median [IQR])** | 10 (5,9) | 13 (13,6) | 9,3 (2,9) | 0.18 |

\*Values are n (%) unless otherwise indicated. Continuous variables are reported as median (IQR). P values from Mann–Whitney U (continuous) or Fisher’s exact test (categorical).

**Table 2.**
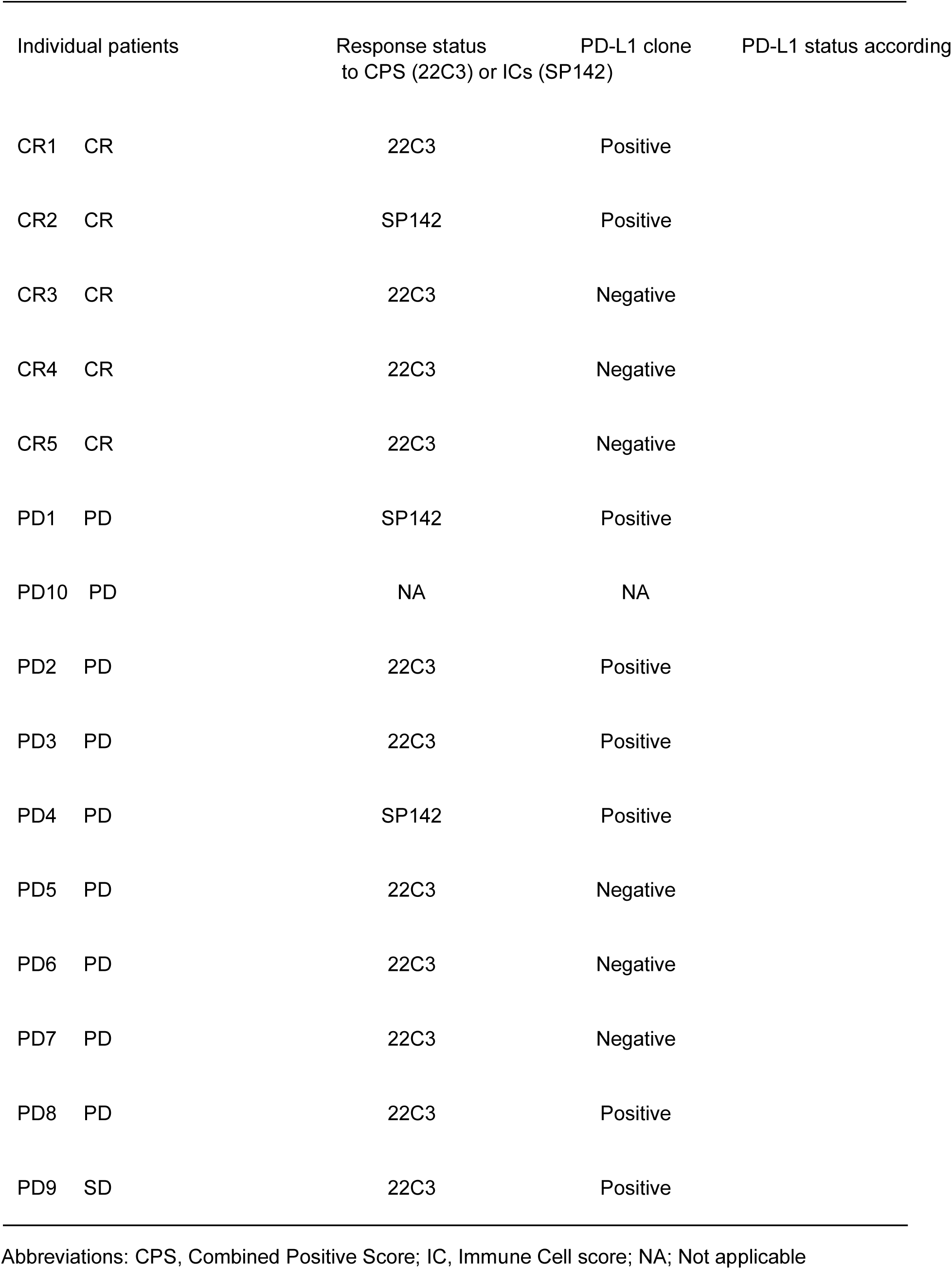
PD-L1 Status, Clinical Immunohistochemistry Diagnostics.

### Comparable immune cell composition in responders and non-responders

All samples contained viable tumor tissue and were annotated into intratumoral and peritumoral regions based on H&E morphology, supported by pan-cytokeratin staining (**Fig. 3A**). Using mIF, immune cell subsets were quantified within these regions, including cytotoxic T cells (CD3⁺ CD8⁺), T helper cells (CD3⁺ CD4⁺ FOXP3-), regulatory T cells (CD3⁺CD4⁺FOXP3⁺), macrophages (CD68⁺), B cells (CD20⁺), with antigen-presenting phenotypes defined by HLA-DR expression across relevant lineages. Representative spatial maps from one CR and one PD case illustrate the annotated immune cell distributions (**Fig. 3B**). Spatial plots for the remaining samples are shown in Supp Fig 2 and also available at this online link: https://is-pla.serve.scilifelab.se for more interactive interpretation.

**Fig. 3.**
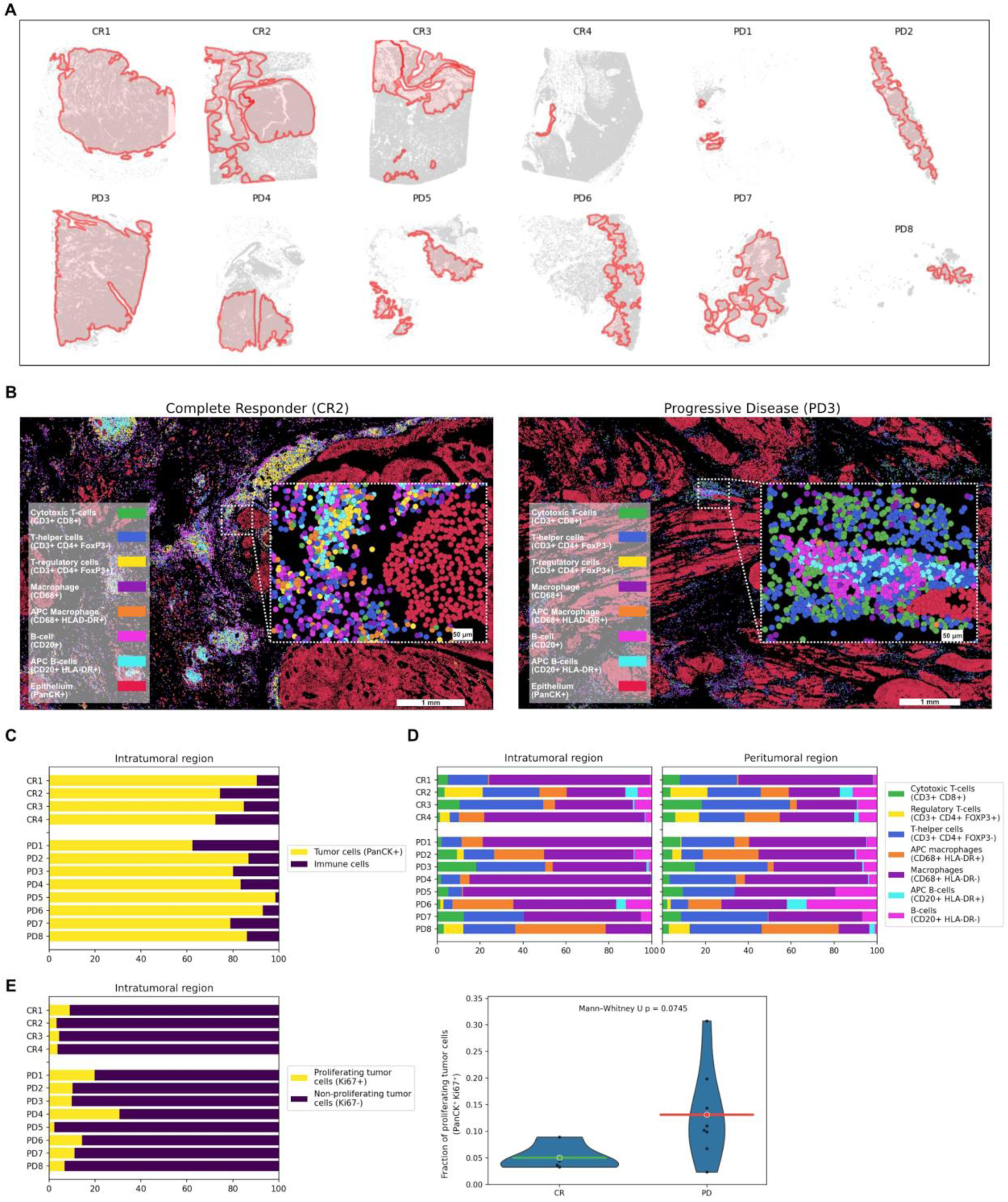
Spatial characterization of tumor and immune cell composition in pembrolizumab-treated urothelial cancer. (A) Whole-slide maps of tumor samples from complete responders (CR1–CR4) and patients with progressive disease (PD1–PD8), with tumor boundaries annotated in red. (B) Representative segmentation overlays from one complete responder (CR2) and one progressive disease case (PD3), showing PanCK⁺ tumor cells (red) and annotated immune cell phenotypes (colored overlays; legend). Insets highlight local tumor–immune organization at single-cell resolution. (C) Quantification of intratumoral tissue composition showing the relative proportions of PanCK⁺ tumor cells and immune cells across individual samples. (D) Relative distribution of annotated immune cell phenotypes within intratumoral and peritumoral regions, stratified by clinical response. (E) Quantification of proliferating (PanCK⁺Ki67⁺) and non-proliferating (PanCK⁺Ki67⁻) tumor cells within intratumoral regions. Violin plots summarize the fraction of proliferating tumor cells per sample. Statistical comparisons were performed using the Mann–Whitney U test.

The fraction of immune cells within intratumoral regions ranged from a few percent to approximately 20% but did not differ significantly between CR and PD patients (**Fig. 3C**). Across samples, macrophages represented the most abundant immune population, followed by CD4 ⁺ T cells, with no significant differences in the relative abundance of any immune cell subset between responders and non-responders, nor between intratumoral and peritumoral regions (**Fig. 3D** and **Supp.Fig. 3).**

Tumor cell proliferation, assessed by Ki-67 expression, varied substantially across samples (1–30%). Although higher proliferation rates were observed in PD compared with CR patients (mean 12% vs. 5%), this difference did not reach statistical significance (Mann Whitney p 0,07) (**Fig. 3E-3F**).

### Single-marker PD-L1 expression across response groups

Using multiplex immunofluorescence (mIF), we quantified single-marker PD-L1 expression within the intratumoral region, independent of PD-1/PD-L1 interaction status. Spatial mapping revealed substantial inter-sample variability in PD-L1 expression, ranging from nearly PD-L1–negative to predominantly PD-L1–positive cases (**Fig. 4A**). PD-L1–positive cells included both tumor cells and immune cells and were heterogeneously distributed across samples in both clinical response groups.

**Fig. 4.**
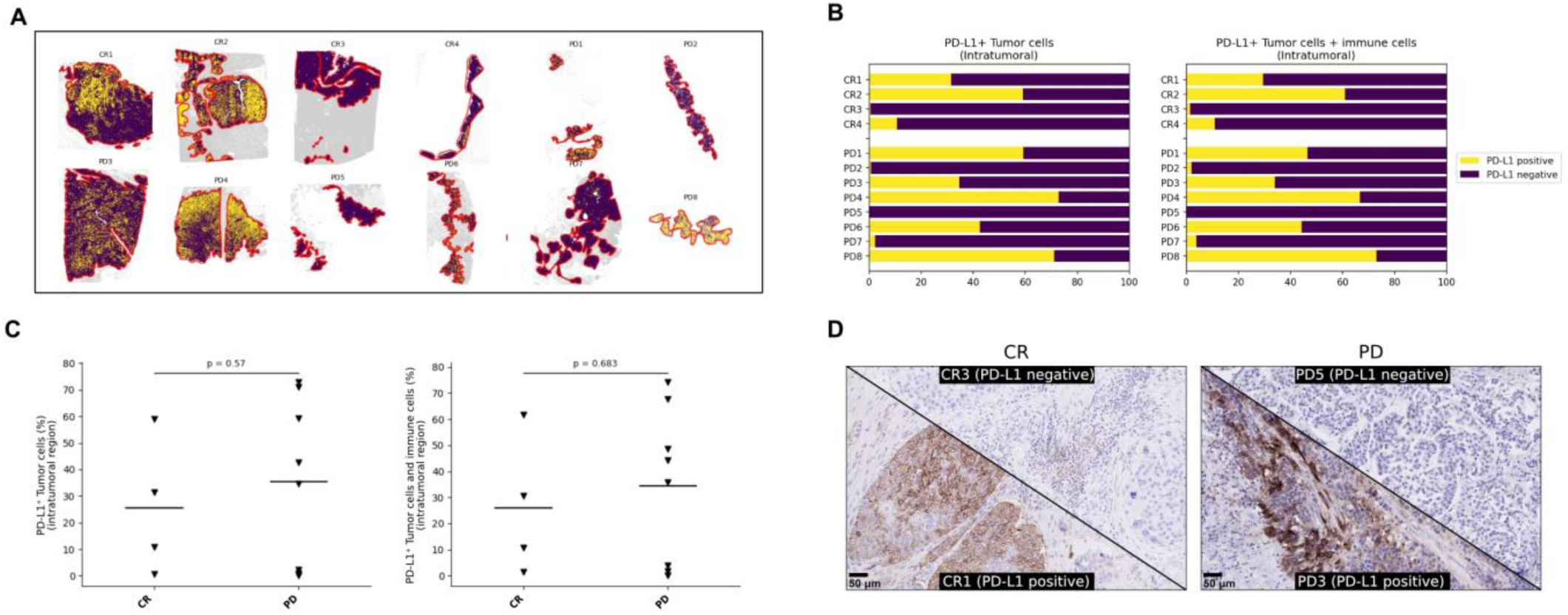
Single-marker PD-L1 expression across clinical response groups. (A,B) Spatial quantification of PD-L1–positive cells within the intratumoral region, including PD-L1⁺ tumor cells alone or combined with PD-L1⁺ mononuclear immune cells, stratified by clinical response. (C). Left, percentage of PD-L1⁺ PanCK⁺ tumor cells relative to all viable PanCK⁺ tumor cells; right, percentage of PD-L1⁺ PanCK⁺ tumor cells combined with PD-L1⁺ mononuclear immune cells relative to the total number of viable PanCK⁺ tumor cells. Each triangle represents one patient (complete responder [CR] or progressive disease [PD]). Statistical comparisons were performed using the Mann–Whitney U test. (D) Representative routine clinical PD-L1 IHC images from PD-L1–negative and PD-L1–positive complete responders (CR3 and CR1) and patients with progressive disease (PD5 and PD3).

In line with clinical PD-L1 assessment practices in urothelial cancer, PD-L1 expression was evaluated as a single marker in tumor cells alone as well as in tumor cells combined with immune cells (**Fig. 4B**). Across the cohort, the fraction of PD-L1–positive cells varied widely, from a few percent to more than 70%, within both complete responder (CR) and progressive disease (PD) groups. No significant differences in PD-L1 positivity were observed between CR and PD patients when assessed either in tumor cells alone or in the combined tumor and immune cell compartment (**Fig. 4C**).

PD-L1 expression levels measured by mIF were concordant with PD-L1 status assessed by routine clinical immunohistochemistry (IHC) (**Table 2**). Representative examples of PD-L1–positive and PD-L1–negative clinical IHC staining from CR and PD patients are shown in **Fig. 4D**. Collectively, **Fig. 4** demonstrates that single-marker PD-L1 expression, assessed by both mIF and routine IHC, does not distinguish clinical response groups in this cohort.

### PD-1/PD-L1 interactions are enriched in the peritumoral region of complete responders

We next analyzed isPLA signals across samples to quantify PD-1/PD-L1 interactions and assess their association with clinical response. Representative images illustrate regions with dense isPLA signals in a complete responder (CR) sample compared with more sparse signals in a progressive disease (PD) sample (**Fig. 5A**).

**Fig. 5.**
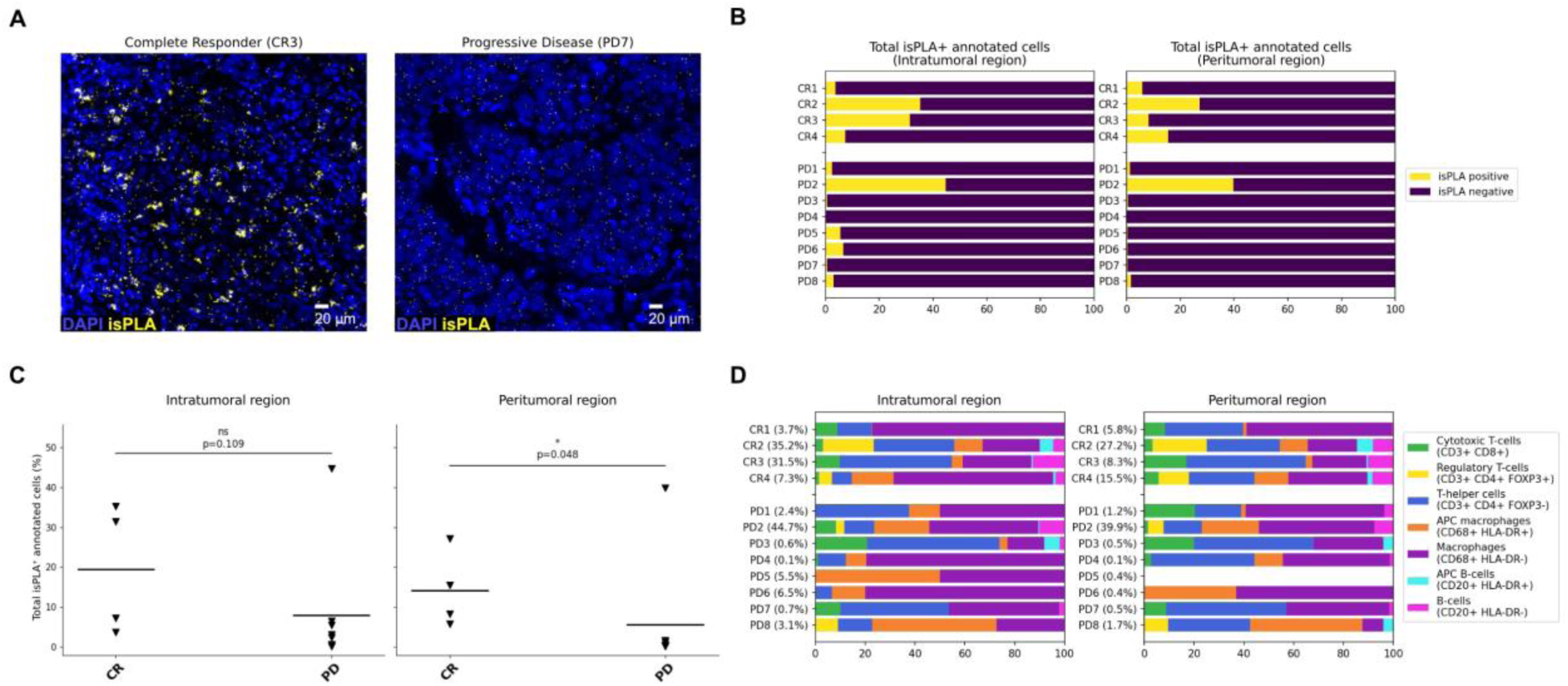
Spatial quantification of PD-1/PD-L1 interactions by isPLA. (A) Representative isPLA images from a complete responder (CR3, left) and a patient with progressive disease (PD7, right), illustrating the spatial distribution of isPLA signals. (B) Fraction of isPLA-positive cells per analyzed sample (CR1–CR4, PD1–PD8) within intratumoral and peritumoral regions. Bars represent the proportion of isPLA-positive and isPLA-negative cells, with all annotated cells per sample normalized to 100%.n(C) Comparison of the fraction of isPLA-positive cells between complete responders (CR) and patients with progressive disease (PD) within intratumoral and peritumoral regions. Statistical comparisons were performed using the Mann–Whitney U test. (D) Relative distribution of isPLA-positive cells across annotated cellular phenotypes for each analyzed sample (CR1–CR4, PD1–PD8). The percentage of total isPLA-positive cells in respective sample is indicated in parentheses next to each sample identifier.

To enable quantitative comparisons across samples, isPLA signals detected in the DO channel were assigned to individual cells, and cells displaying at least one isPLA spot were classified as isPLA-positive. The fraction of isPLA-positive cells among all annotated cells was quantified separately in intratumoral and peritumoral regions (**Fig. 5B**). Across samples, the fraction of isPLA-positive cells varied within both response groups, with a consistent trend toward higher fractions in CR patients. Notably, higher fractions of isPLA-positive cells were generally observed in peritumoral regions. In several PD cases, isPLA-positive cells were absent from the intratumoral compartment but present in peritumoral regions.

Comparison between response groups revealed no statistically significant difference in the fraction of isPLA-positive cells within the intratumoral region, although a trend toward higher values was observed in CR patients (**Fig. 5C**). In contrast, the fraction of isPLA-positive cells in the peritumoral region was significantly higher in CR compared with PD patients (**Fig. 5C**).

To further characterize the cellular context of PD-1/PD-L1 signaling, isPLA-positive cells were assigned to annotated cell phenotypes (**Fig. 5D**). isPLA signals were detected across all major immune cell types, although their relative abundance varied between samples. CD4⁺ T cells and CD68⁺ macrophages (HLA-DR⁺/⁻) represented the dominant isPLA-positive populations across both intratumoral and peritumoral regions in all samples. In contrast, isPLA-positive B cells (HLA-DR⁺/⁻) and FOXP3⁺ regulatory T cells were infrequent and not detected in all cases. Notably, isPLA-positive cytotoxic CD8⁺ T cells were consistently observed in all CR patients across both regions, whereas such cells were rare in PD patients. While the overall distribution of isPLA-positive cell types was broadly similar between CR and PD patients, the relative contribution of cytotoxic CD8⁺ T cells to PD-1/PD-L1 interactions was increased in CR patients.

### Cytotoxic CD8⁺ T cells drive PD-1/PD-L1 interaction differences between response groups

Building on the cell-type–resolved analysis of isPLA-positive cells (**Fig. 5D**), we performed statistical testing to assess differences between complete responders (CR) and patients with progressive disease (PD) with respect to the cellular contributors to PD-1/PD-L1 interactions. This analysis identified cytotoxic CD3⁺CD8⁺ T cells as the only cell population showing consistently higher fractions of isPLA positivity in CR patients across spatial compartments.

Specifically, the fraction of isPLA-positive cytotoxic CD8⁺ T cells was significantly increased in CR patients within both the peritumoral region (p = 0.013) and the intratumoral region (p = 0.031) and, as well as when regions were analyzed in combination (p = 0.033) (**Fig. 6**). In contrast, other immune cell types showed more region-restricted associations. isPLA-positive CD4⁺ T cells, macrophages, and B cells were significantly enriched in CR patients only within the peritumoral region, while no significant differences were observed intratumorally or in combined analyses.

**Fig. 6.**
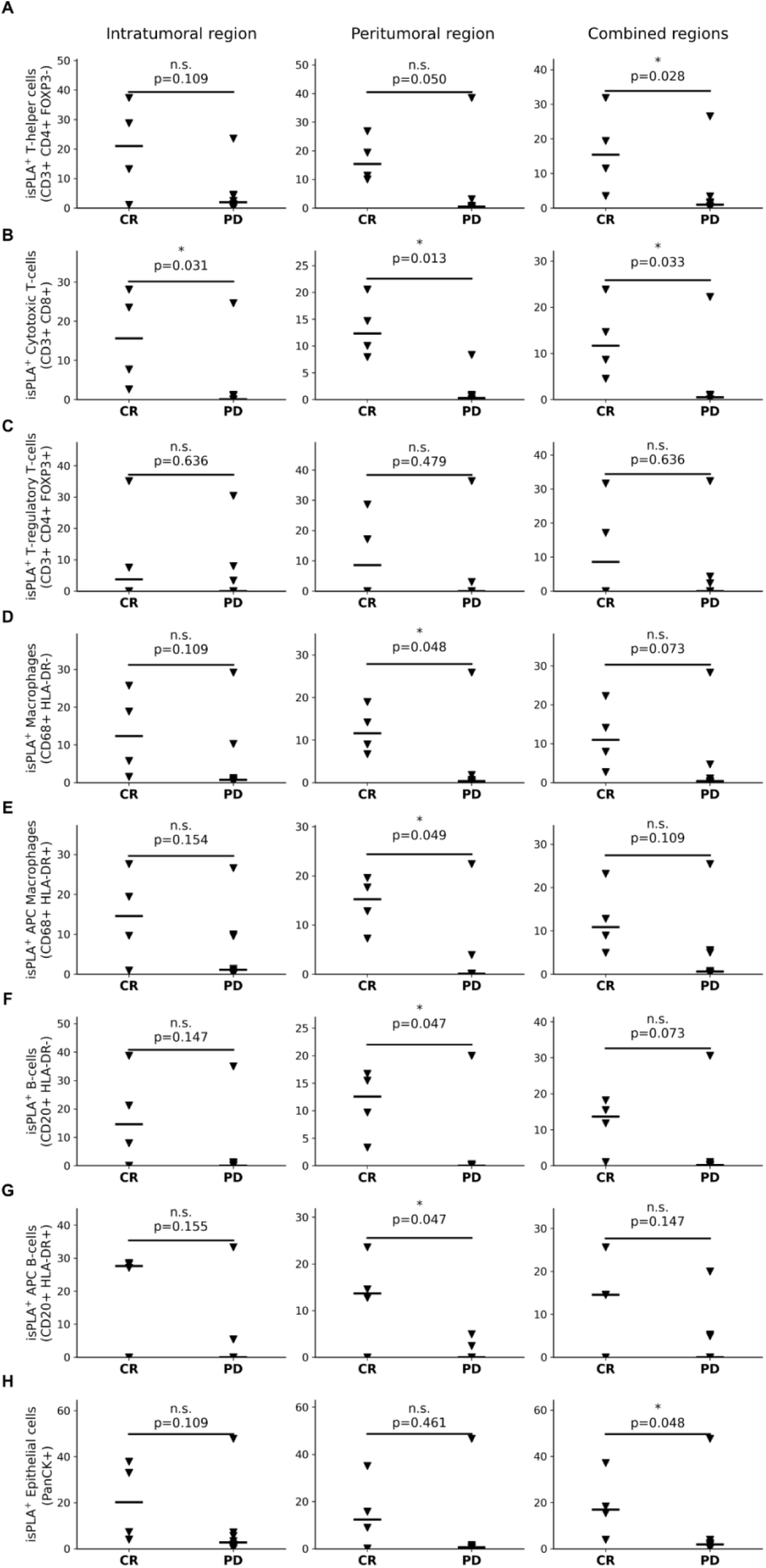
Cell-type–resolved quantification of PD-1/PD-L1 interactions across tumor regions. Fraction of isPLA-positive annotated cell types, quantified separately within the intratumoral region, peritumoral region, and across the combined regions. Each data point represents one patient (complete responder [CR] or progressive disease [PD]). Statistical comparisons were performed using the Mann–Whitney U test.

Importantly, our earlier analysis of immune cell composition demonstrated the presence of cytotoxic T cells in tumors from both CR and PD patients. The integration of isPLA data therefore provides functional resolution, revealing that a substantially higher proportion of these cytotoxic T cells actively engage in PD-1/PD-L1 signaling in CR patients. The consistency of this association across intratumoral, peritumoral, and combined regions distinguishes cytotoxic CD8⁺ T cells from other immune populations and identifies them as the dominant cellular correlate of PD-1/PD-L1 interaction differences between response groups.

*isPLA-positive cytotoxic CD8⁺ T cells define an interaction-based biomarker signature* Building on the consistent enrichment of PD-1/PD-L1 interactions in cytotoxic CD8⁺ T cells across tumor regions (**Fig. 6**), we next examined the biomarker potential and functional state of this interaction-defined T-cell subset. Representative images illustrate that both complete responders (CR) and patients with progressive disease (PD) exhibit infiltration of CD3⁺CD8⁺ cytotoxic T cells within tumor regions (**Fig. 7A**). However, PD-1/PD-L1 signaling between these T cells and tumor cells is observed only in the CR patient (left), while such interaction is absent in the PD sample (right). Voronoi diagrams were generated to explore the spatial context of these events. Colour-coding of tumor cells, immune cells, and isPLA⁺ cytotoxic T cells highlight that isPLA-positive CD3⁺CD8⁺ T cells penetrate the intratumoral regions in CR patients, whereas such cells are rare or lacking in PD patients (**Fig 7B**, **Supp fig 4**).

**Fig. 7.**
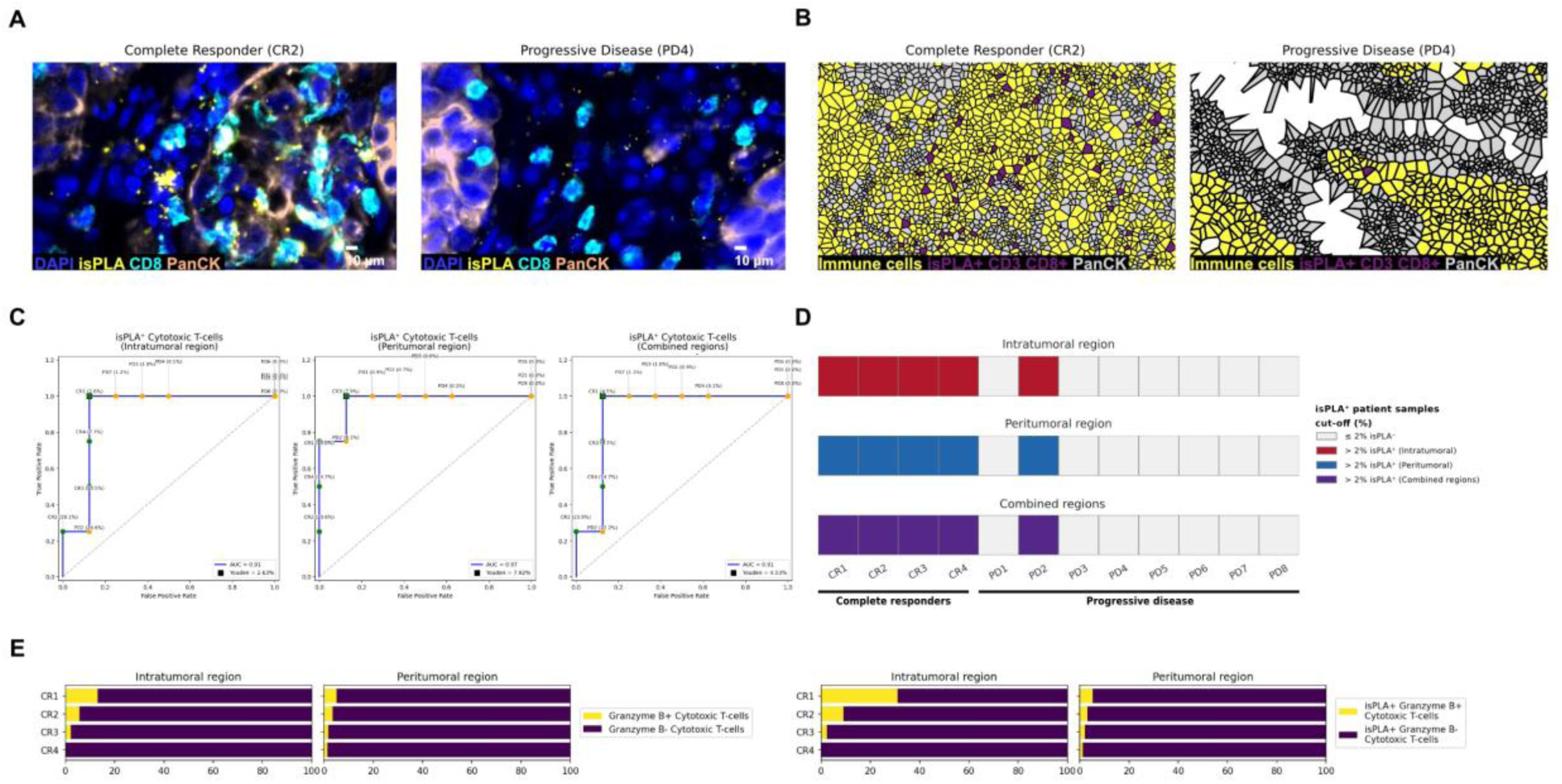
Visualization and characterization of interaction-defined cytotoxic T cells. (A) Representative multiplexed images showing cytotoxic T cells (CD3⁺CD8⁺, cyan), PanCK⁺ tumor cells (orange), and PD-1/PD-L1 interactions detected by isPLA (yellow) in tumor regions. Nuclei are counterstained with DAPI (blue). (B) Voronoi diagrams of representative intratumoral regions illustrating the spatial distribution of tumor cells (grey), immune cells (yellow), and isPLA-positive cytotoxic T cells (blue).(C) Receiver operating characteristic (ROC) analysis evaluating the performance of isPLA-positive cytotoxic T cells as a binary classifier of clinical response, based on intratumoral, peritumoral, and combined regions. (D) Classification of individual patient samples using a fixed cutoff for the fraction of isPLA-positive cytotoxic T cells, applied across intratumoral, peritumoral, and combined regions. (E) Quantification of Granzyme B expression in cytotoxic T cells, comparing the total CD3⁺CD8⁺ population with the isPLA-positive subset.

We next evaluated the performance of isPLA-positive cytotoxic CD8⁺ T cells as a binary biomarker for separating CR from PD patients using receiver operating characteristic (ROC) analysis. Using cutoffs for the fraction of isPLA-positive CD3+ CD8+ cells as part of all CD3+CD8+ cells, we could clearly separate all our CR patients from the PD group with an area under the curve (AUC) above 0,9 for both intratumoral (cutoff 2%, AUC = 0,91), peritumoral (cutoff 7 %, AUC = 0,97) or combined areas (cutoff 4 %, AUC = 0,91) (**Fig 7C**). From a more pragmatic clinical perspective, using a cutoff of 2 % allowed us to separate CR from PD patients independently of annotated intratumoral and peritumoral areas (**Fig 7D**). Patient PD2 showed to be an exception by displaying a fraction of isPLA positive cytotoxic T cells above 2%. This sample, unlike the others obtained at primary diagnosis, was derived from a metastatic core biopsy taken at a later disease stage, after only one cycle of pembrolizumab and has appeared as an outlier in multiple analyses throughout the study.

Given the limited predictive value of PD-L1 expression alone, we compared the performance of this interaction-based biomarker with single-marker approaches. PD-L1 positivity assessed by mIF showed no discriminatory power between CR and PD patients (AUC = 0.5), consistent with routine clinical IHC results (**Supplementary Fig. 5**). Similarly, neither total isPLA-positive cells nor CD3⁺CD8⁺ T cells alone achieved comparable classification performance. These comparisons demonstrate that the combined information provided by PD-1/PD-L1 interaction status and cytotoxic T-cell identity outperforms single-marker biomarkers in separating responders from non-responders.

Lastly, we assessed cytolytic activity among cytotoxic T cells using Granzyme B as a functional marker. When comparing all cytotoxic T cells to the isPLA-positive subset, we observed a clear enrichment of Granzyme B⁺ cells within the isPLA-positive population (left chart), indicating that these cells are more functionally active (**Fig 7E**). These findings suggest that a subset of the T cells interacting with tumor cells are functionally active and participate in tumor cell elimination. As expected, we also observed higher proportions of activated cytotoxic T cells within the intratumoral compared to the peritumoral region. This analysis underscores the strength of combining the isPLA assay, which identifies cell–cell interactions, with phenotypic and functional markers from mIF, thereby providing higher spatial resolution and deeper biological insight.

## DISCUSSION

In this work, we have developed an integrated automated assay that combines isPLA with mIF, to directly link PD-1/PD-L1 signaling events to defined cellular phenotypes and their spatial organization within intact tumor tissue. Applied, as a proof-of-concept, to samples from patients with advanced urothelial carcinoma treated with pembrolizumab, this approach reveals that functional checkpoint engagement -specifically between cytotoxic CD8⁺ T cells and tumor cells - provides substantially stronger discrimination of clinical response than PD-L1 expression alone.

A central strength of the platform is the generation of a single, fully integrated dataset in which protein–protein interactions and high-dimensional phenotyping are acquired from the same tissue section and annotated at the level of individual cells. This avoids the need for post image co-registration and ensures that signaling events are unambiguously assigned to their cellular source and spatial niche. The resulting interaction-resolved maps move beyond proximity-based inference and provide direct evidence of functional immune checkpoint engagement within the tumor microenvironment.

Our findings demonstrate that complete responders are characterized by a consistent enrichment of PD-1/PD-L1 interactions involving cytotoxic CD8⁺ T cells across both intratumoral and peritumoral compartments. This observation refines current models of ICI responsiveness by indicating that the mere presence of effector T cells is insufficient; rather, it is the active engagement of these cells in checkpoint signaling that marks a tumor as immunologically poised to respond to PD-1 blockade. The enrichment of Granzyme B within the isPLA-positive CD8⁺ T-cell subset further supports the interpretation that these interaction-defined cells represent a functionally active, cytolytic population directly involved in antitumor immunity. These data are consistent with studies in melanoma showing that tumor-infiltrating CD8⁺ T cells with defined phenotypic and functional properties correlate with antitumor activity and clinical outcomes (20). The spatial distribution of signaling events also highlights the biological relevance of the peritumoral microenvironment. While intratumoral interactions predominantly reflect direct tumor–T-cell engagement, the peritumoral compartment in responders shows broader participation of immune subsets, including CD4⁺ T cells, macrophages, and B cells. This pattern suggests a coordinated immune network extending beyond the tumor core and is consistent with emerging evidence that stromal and immune niches surrounding the tumor can strongly influence therapeutic outcomes. The parallel with recent spatial multi-omics studies in non–small-cell lung cancer, which link favorable immunotherapy responses to immune-rich stromal signatures, underscores the general importance of extratumoral immune organization as a determinant of treatment efficacy (21). In addition, our results complement prior work on T-cell–inflamed and immune-excluded tumor phenotypes as determinants of checkpoint inhibitor efficacy, providing a direct, interaction-based readout of these immune states (22).

By contrast, single-marker PD-L1 expression-assessed by both multiplexed IF and routine clinical immunohistochemistry-showed no discriminatory power in this cohort. This aligns with prior reports documenting the limited and context-dependent predictive value of PD-L1 staining and reinforces the conceptual limitation of using static expression readouts to predict the efficacy of therapies that target dynamic, cell–cell signaling processes. The superior performance of an interaction-based, cell-type–resolved biomarker emphasizes the value of measuring functional immune engagement rather than surrogate markers of pathway availability.

From a translational perspective, the present workflow demonstrates how highly multiplexed, interaction-resolved imaging can be adapted to clinically relevant platforms and analysis pipelines. Although the full 14-marker panel provides deep phenotypic resolution, our data indicate that robust response prediction can be achieved using a minimal marker set comprising PD-1/PD-L1 interaction detection together with CD3 and CD8. This reduction in complexity opens a path toward more cost-effective, scalable implementations compatible with standard autostainers and digital pathology infrastructure, potentially facilitating clinical translation.

Several limitations should be considered, consistent with the proof-of-concept nature of this study. The cohort was intentionally enriched for patients at the extremes of clinical response to maximize contrast in immune signaling phenotypes, and future studies in larger, unselected populations will be important to assess the robustness and generalizability of the interaction-based biomarker. Despite these constraints, in contrast to many biomarker studies that rely on tissue microarrays, our use of whole-tissue sections provides a more comprehensive representation of tumor–immune architecture and enables interrogation of peritumoral regions that are often excluded from high-throughput approaches. Furthermore, when the aim is to identify predictive biomarkers relevant for real-time pathological evaluation, analysis of whole-tissue sections aligns more closely with routine clinical workflows and the manner in which diagnostic specimens are assessed in practice.

In summary, this work establishes a framework for integrating direct detection of protein–protein interactions with spatially resolved, single-cell phenotyping to derive mechanistically informed biomarkers of immunotherapy response. In metastatic urothelial carcinoma, PD-1/PD-L1 engagement by cytotoxic T cells emerges as a putative dominant and functionally meaningful proxy of pembrolizumab efficacy. More broadly, the approach is adaptable to other receptor–ligand systems and tumor types, providing a generalizable strategy for translating spatial signaling biology into predictive tools for precision oncology.

## MATERIALS AND METHODS

### Study Design

The overall objective of the study was to explore PD-1/PD-L1 signaling between defined cell types as a biomarker for response to pembrolizumab immunotherapy and further compare its performance with current PD-L1 staining as a biomarker in urothelial carcinoma. A substantial component of the study involved the development and integration of analytical methods and workflows to enable cell type–resolved assessment of PD-1/PD-L1 signaling on the commercial PhenoCycler platform.

The study was conducted on a subset of surgical specimens from a larger retrospective cohort of all mUC patients treated with pembrolizumab between the years of 2017-2022 at the Karolinska University hospital (n = 107). The cohort is well characterized and has previously been described (23). For the present proof of concept study, a sub cohort of 15 patient cases were selected based on treatment response; 5 patients with complete response (CR) and 10 non-responders with progressive disease (PD). The rationale for selecting these specific cases was twofold: first, to capture patients representing opposite ends of the ICI response spectrum and thereby maximize the likelihood of detecting immunological differences between tumors; and second, to include variation in PD-L1 status, allowing comparison of the current clinical gold standard biomarker for ICI therapy with our isPLA- and cell phenotyping–based biomarker approach.

### Human tissue samples

Formalin-Fixed Paraffin-Embedded (FFPE) was collected from primary tumors for all patients, except in one case where a sample from a distant metastasis was used due to the unavailability of primary tumor tissue. Due to insufficient material from two of the fifteen patients, a total of thirteen FFPE material tumor samples were included in the final analysis. The clinical characteristics and pathology data for each group are summarized in **Table 1**. All patient samples were collected and used in accordance with ethical approval (permit number: Dnr: 2022-0164501). For the quality control runs, bladder cancer tissue was retrieved from anonymized residual material retrieved from the pathology archive.

### Tissue preparations for standard PhenoCycler runs

Urothelial cancer and tonsil tissue used for isPLA assay optimization and validation were processed according to Akoya Bioscience’s (Quanterix) standard protocol for dewaxing, deparaffinization/rehydration and tissue staining of FFPE tissue samples and as previously described (24,25).

### Tissue preparation for detection of PD1/PD-L1 interaction using isPLA

All clinical UC FFPE samples underwent tissue processing and PD-1/PD-L1 isPLA staining using Naveni^Ⓡ^ PD-1/PD-L1 Atto647N kit (art. no. 60011) on Leica Bond RX autostainer (Leica, UK) at Navinci Diagnostics facilities (Uppsala). A prototype of the now commercially available protocol was executed according to the instructions for PD-1/PD-L1 isPLA detection (26). The commercial protocol now includes additional washing steps post navenibody-staining step (including two 20 min washes at 37 ℃), for improved staining specificity and reduced background.

### Integrated isPLA and PhenoCycler data generation

The tissue sections were pre-processed according to our standard procedure for the PhenoCycler assay, followed by the automated isPLA protocol developed for Leica Bond as described above. Sections were subsequently incubated with the full antibody panel for the PhenoCycler assay, as detailed above. The slides were put into the flow cells for image data capture across cycles on the PhenoCycler Fusion. The reporter plate containing the fluorescently labeled reporters for both the isPLA product and the corresponding reporters for the barcoded antibodies was prepared according to standard PhenoCycler runs, with the detection oligo for isPLA product placed separately for detection in the first imaging cycle.

In detail, the Naveni Detection Mix (5x) Atto647n was diluted 5x and mixed with Akoya Bioscience’s reporter buffer assay. The solution was then added to the first well of a 96-well reporter plate. Following this, a blank reporter buffer assay (excluding reporters) was added to the second well in order for the chemical de-attachment of the detection oligos from the PD-1/PD-L1 rolling-circle-tree. Finally, reporters for a 14-plex panel were dispensed in the remaining wells for multiplex phenotyping of cell markers; CD3, CD4, CD8, CD20, C31, CD68, HLA-DR, Pan-cytokeratin, FoxP3, PD-1, PD-L1, Granzyme B, PCNA and Ki67.

### Image data analysis and visualization

Imaging data were exported as 8-bit compressed qptiff files and processed using the PIPEX software pipeline. Cell segmentation was performed with StarDist (default parameters; nuclei diameter = 20 pixels, cytoplasmic expansion = 20 pixels), followed by extraction of mean fluorescence intensities for each marker per segmented cell.

Detection of isPLA signal was carried out using the BigFish library. Images were denoised and filtered with a Laplacian of Gaussian (LoG) filter to enhance isPLA spots, after which local maxima were identified. An adaptive intensity threshold was applied to distinguish true spots from background noise. Dense and bright regions were further decomposed using Gaussian simulations to correct potential misdetections. For each cell, the total number of isPLA spots was quantified. Cluster detection modules were available but not applied in this study.

The resulting cell data, including marker intensities and isPLA spot counts, were stored in AnnData objects for downstream analysis. Consecutive H&E-stained tissue sections were manually aligned to the DAPI channel images to guide pathologist annotations (TissUUmaps v. 3.0). Pathologist-curated annotations were used to delineate intratumoral versus peritumoral regions, with each cell labeled accordingly.

Quality control of isPLA signal was performed in collaboration with an isPLA assay expert from Navinci Diagnostics, who systematically annotated and flagged regions affected by artifacts, edge effects or technical noise. This step ensured that only high-confidence isPLA signal was included in downstream analysis.

For marker positivity, Otsu thresholding (or multi-Otsu for three-class separation) was applied, with the highest intensity class retained as positive. Multi-Otsu was specifically used for FOXP3, PD-1 and PD-L). isPLA signals were initially categorized into four classes based on per-cell spot counts (zero, low, intermediate and high). However, for the final analysis, a simplified binary threshold was adopted: cells with ≥1 isPLA spots were considered positive and thus capable of forming an interaction with another cell.

Cells were phenotyped using a supervised classification approach (hierarchical binary decision-tree), whereby sequential marker threshold holds defined cell types. Tree leaves representing biologically relevant phenotypes were retained, while others were categorized as “negative” and “ignored” and were excluded from the analysis.

All analysis of the integrated data were performed within intratumoral and peritumoral regions respectively, annotated from H&E stainings by a certified pathologist. All annotated AnnData objects can be visualized in TissUUmaps for interactive spatial exploration at https://is-pla.serve.scilifelab.se.

All custom code used for image processing and downstream analysis has been made publicly available. The PIPEX-based image processing pipeline, including isPLA spot detection using the BigFish library, can be found at https://github.com/BIIFSweden/pipex_bigfish. Jupyter notebooks for downstream analysis, generation of TissUUmaps projects, and figure creation are available at https://github.com/BIIFSweden/6870_isPLA.

### Statistical Analysis

All analyses were conducted using Python version 3.11.8 within a Jupyter Notebook environment (version 4.4.2). Data preprocessing and visualization were performed using pandas (version 2.2.1), NumPy (version 1.26.4), Matplotlib (version 3.8.4), and Seaborn (version 0.13.2).

Comparisons of cell fractions and marker expression between complete responders and progressive disease patient groups were carried out using the Mann–Whitney U test implemented in SciPy (version 1.13.0), as the data did not meet normality assumptions. Predictive performance was assessed using receiver operating characteristic (ROC) curve analysis with scikit-learn (version 1.4.1.post1). A Youden Index was calculated from ROC curves to determine optimal cut-off values for discriminating between complete responders and progressive disease patients.

Voronoi tessellation was applied using SciPy (version 1.13.0) to transform tissue space into polygonal regions, allowing visualization of isPLA^+^ CD8+ Cytotoxic T cells. Polygon size filtering was used to exclude extreme outliers or border artifacts. Subsampling of PanCK+ epithelial and background cells was applied to improve computational efficiency.

All statistical tests were two-sided. Exact p-values are reported, and statistical significance was defined as p < 0.05.

## Data Availability

All data produced in the present work are contained in the manuscript or as material available online.

https://is-pla.serve.scilifelab.se.

https://github.com/BIIFSweden/6870_isPLA.

## Acknowledgments

We acknowledge the Spatial Proteomics Unit at SciLifeLab for providing access to instrumentation. We acknowledge Navinci Diagnostics for providing access to instrumentation and helping with isPLA control experiments and set-up.

## Author contributions

Conceptualization: DK, KH, AU, CS

Methodology: TU, DK, ML, CA

Investigation: TU, CA, DK, FCS, KH, PÖ, AU, CS

Visualization: TU, CA, CS, DK

Funding acquisition: AU, CS

Project administration: AU, CS

Supervision: PÖ, AU, CS

Writing – original draft: TU, CA, CS

Writing – review & editing: TU, CA, ML, KH, FCS, PÖ, DK, AU, CS

## Data and materials availability

All data, code, and materials used in the analysis must be available in some form to any researcher for purposes of reproducing or extending the analysis. Include a note explaining any restrictions on materials, such as materials transfer agreements (MTAs). Note accession numbers to any data relating to the paper and deposited in a public database; include a brief description of the data set or model with the number. If all data are in the paper and supplementary materials, include the sentence “All data are available in the main text or the supplementary materials.

---

**Supplementary Figure 1.**
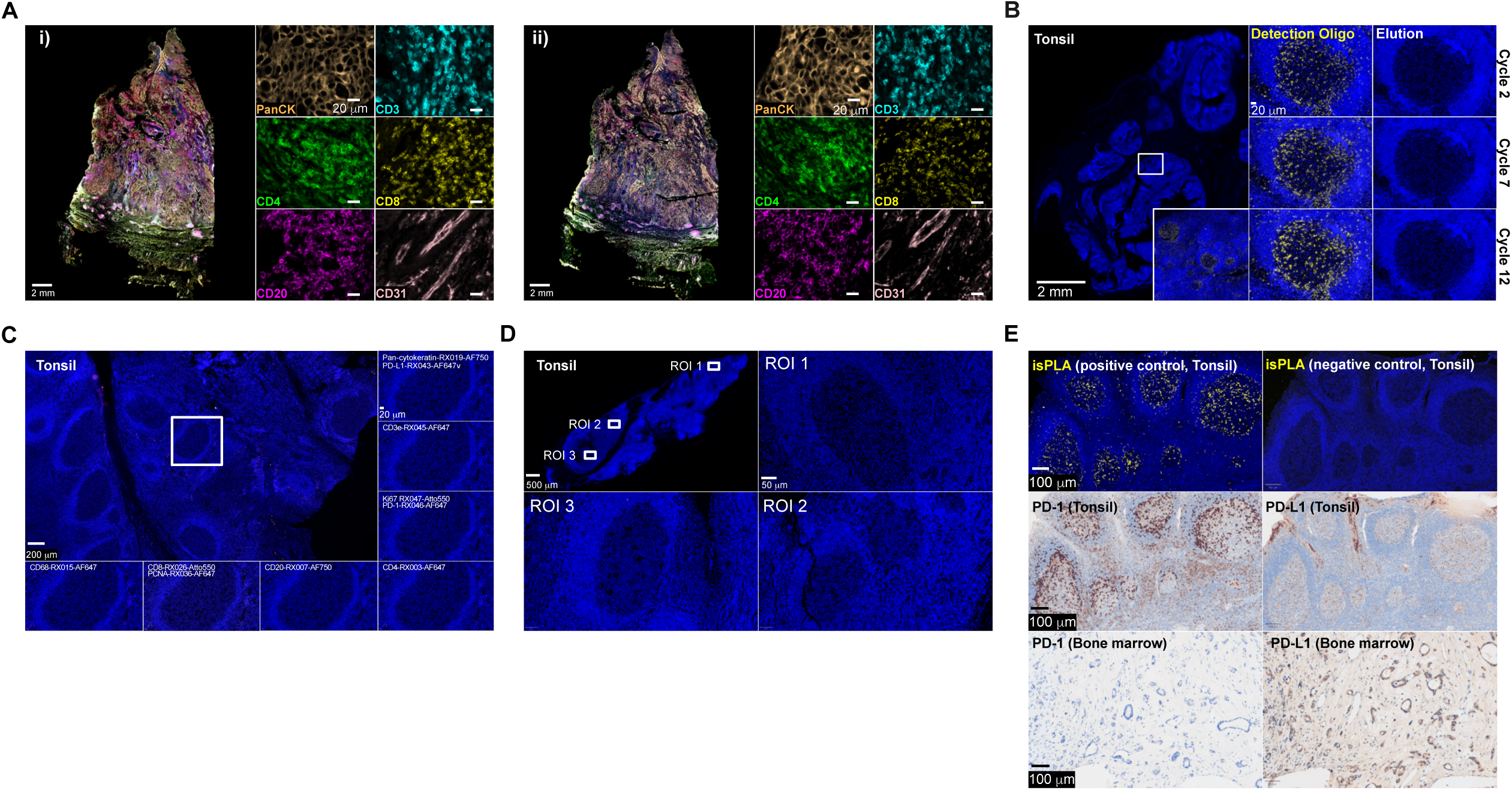
Validation of assay integration and specificity. (A) (i) Urothelial cancer section processed with PD-1/PD-L1 isPLA prior to the PhenoCycler workflow. Subsequent multiplex antibody staining (PanCK, CD3, CD4, CD8, CD20 and CD31) demonstrates preserved marker detection following upstream isPLA. (ii) Control sections processed with the PhenoCycler assay alone are shown for comparison. (B) Tonsil tissue positive control showing PD-1/PD-L1 isPLA, signal stability and reporter elution across imaging cycles (cycles 2, 7 and 12). (C) A cross-reactivity control in tonsil tissue stained with isPLA primary antibodies followed by addition of PhenoCycler reporters, demonstrating absence of assay cross-hybridization. (D) Reciprocal cross-reactivity control in tonsil tissue stained with PhenoCycler antibodies followed by isPLA detection oligonucleotides, confirming lack of nonspecific interactions. (E) Biological assay controls. Tonsil germinal centers showing positive PD-1/PD-L1 isPLA signal and negative controls (no primary antibodies). Chromogenic immunohistochemistry performed as single stains for PD-1 and PD-L1 on consecutive sections confirmed corresponding protein expression. Bone marrow served as a negative tissue control.

**Supplementary Figure 2.**
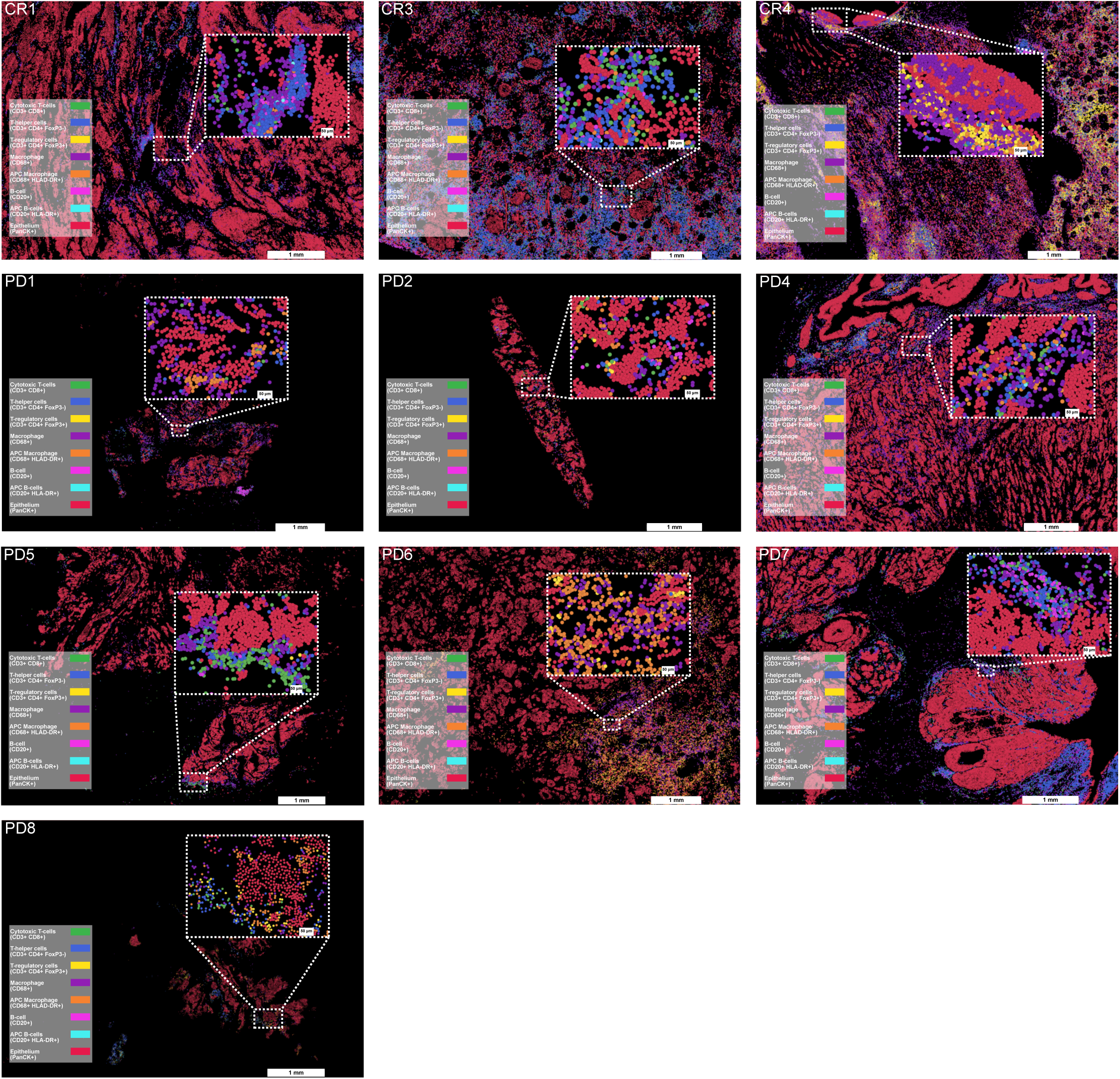
Baseline spatial distribution of major cell phenotypes. Representative urothelial cancer tissue sections from complete responders patients with progressive disease annotated by single-cell segmentation. Spatial maps display cytotoxic T cells (CD3⁺CD8⁺), T-helper cells (CD3⁺CD4⁺FOXP3⁻), regulatory T cells (CD3⁺CD4⁺FOXP3⁺), macrophages (CD68⁺), antigen-presenting macrophages (CD68⁺HLA-DR⁺), B cells (CD20⁺), antigen-presenting B cells (CD20⁺HLA-DR⁺) and epithelial cells (PanCK⁺). Whole-slide views with corresponding high-magnification insets illustrate tissue architecture and immune–tumor organization across response groups.

**Supplementary Figure 3.**
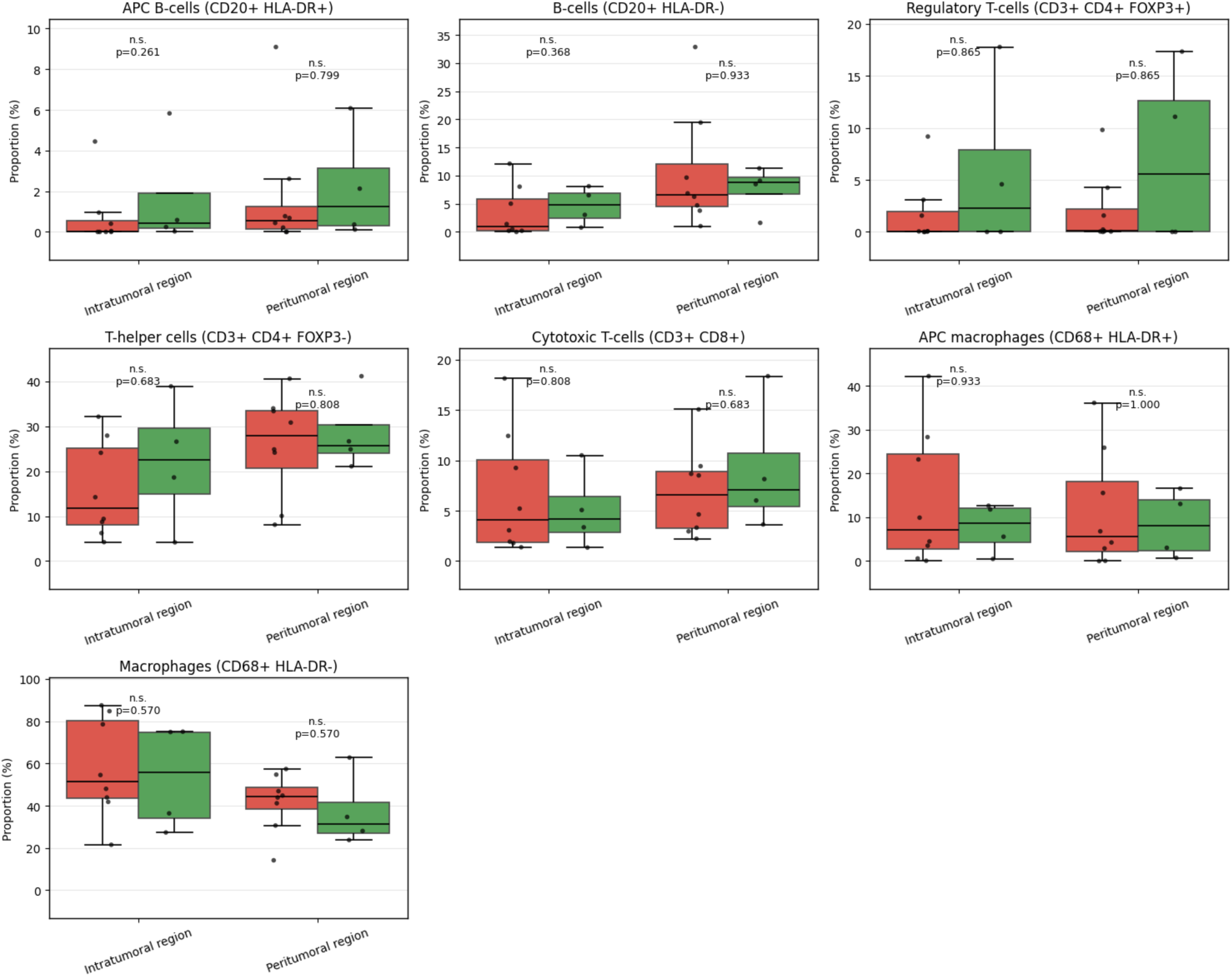
Immune cell composition across patient samples. Boxplots showing relative proportions of major immune cell phenotypes derived from single-cell segmentation of the multiplex dataset. Cell classes include cytotoxic T cells, T-helper cells, regulatory T cells, macrophages, antigen-presenting macrophages, B cells and antigen-presenting B cells. Comparisons are shown between complete responders (grren boxes) and progressive disease cases (red boxes) across intra-and peritumoral regions.

**Supplementary Figure 4.**
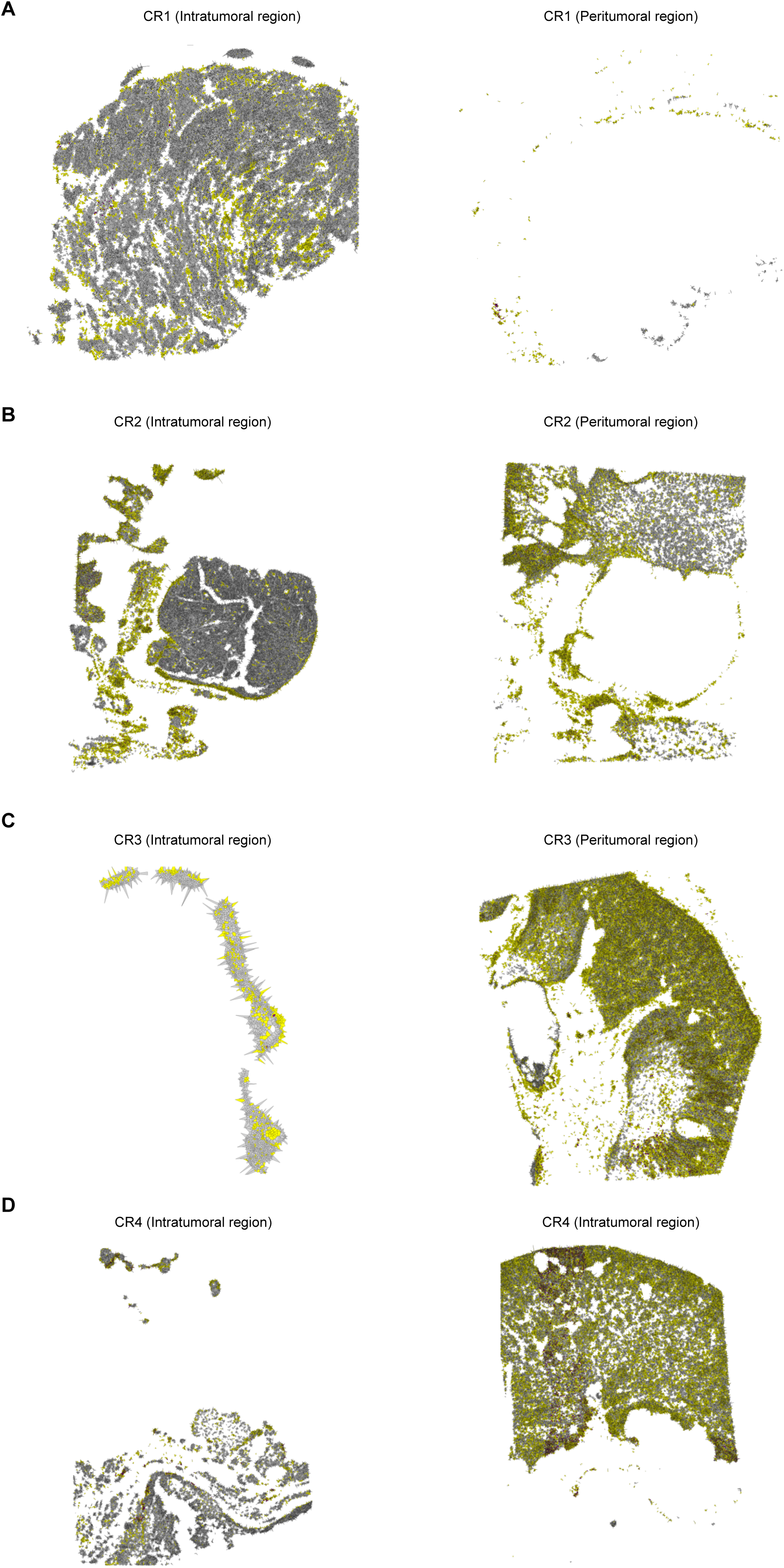
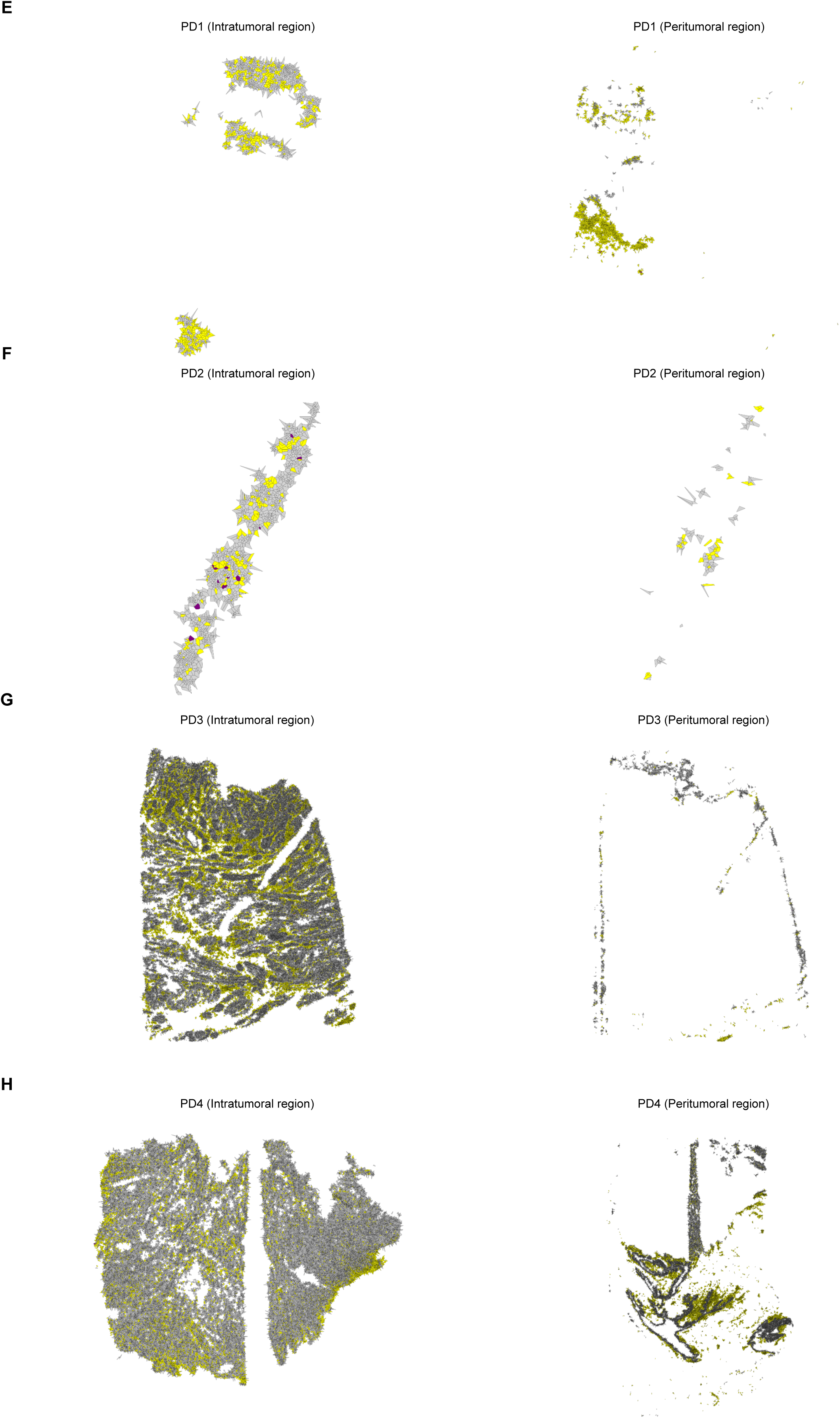
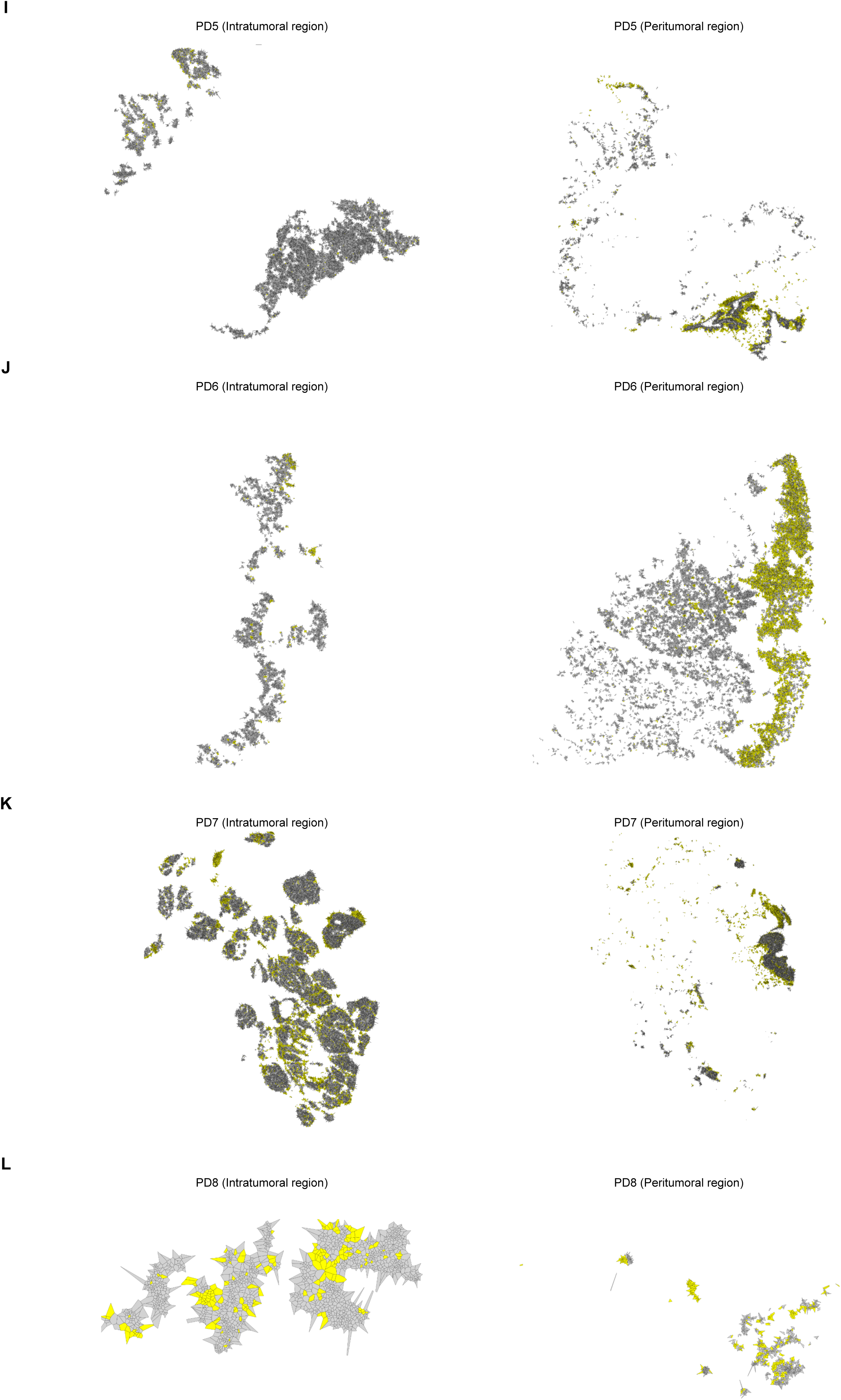
Spatial distribution of PD-1/PD-L1-interacting cytotoxic T cells. Voronoi diagrams illustrating the spatial organization of PD-1/PD-L1 isPLA-positive cytotoxic T cells (purple), other immune cells (yellow), and tumor cells (grey) across patient samples. Diagrams include intra-and peritumoral regions from complete responders and progressive disease cases.

**Supplementary Figure 5.**
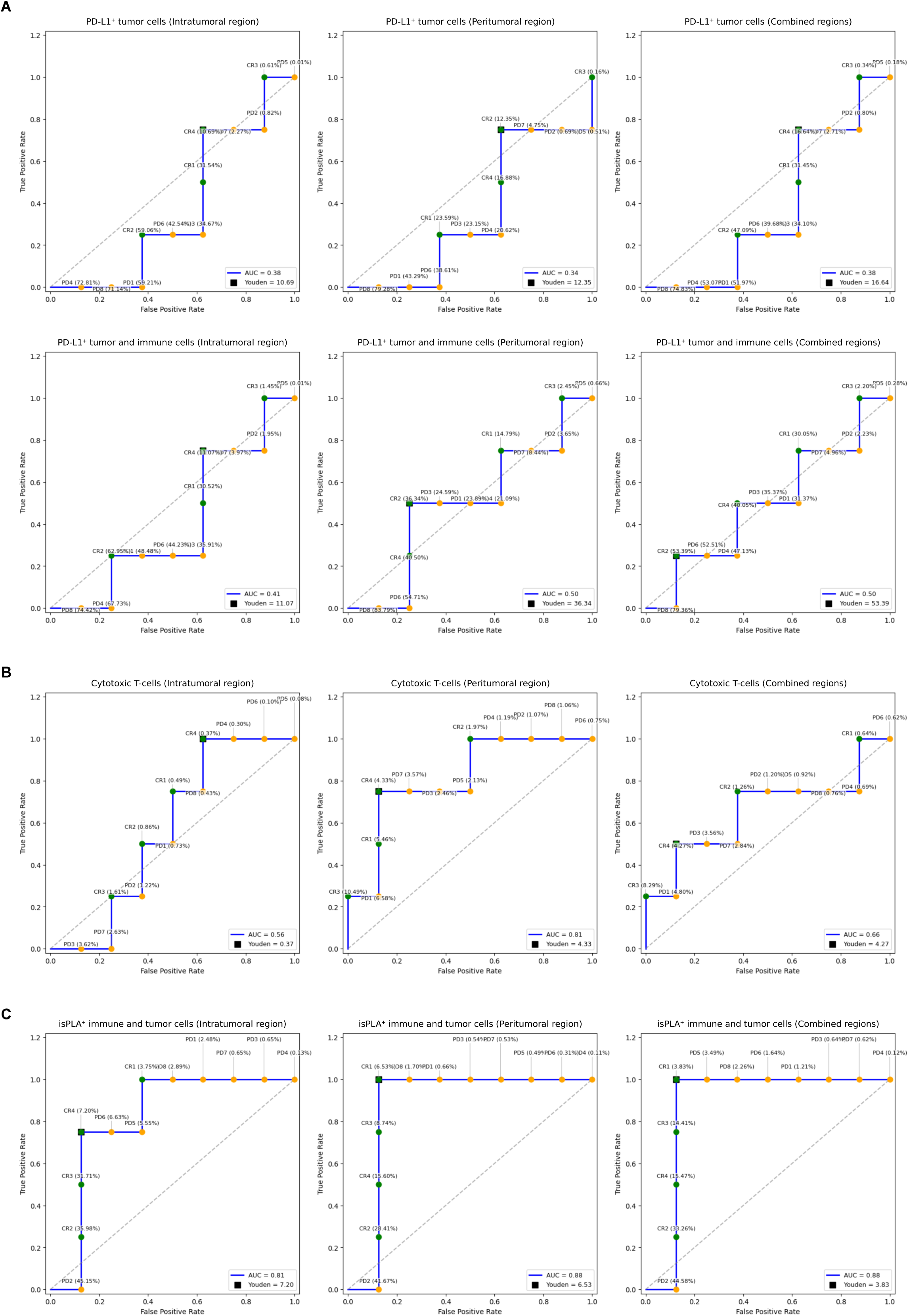
Biomarker performance for response prediction. Receiver operating characteristic (ROC) analyses evaluating response prediction using: A) PD-L1 expression, B) Cytotoxic T-cell abundance, and C) PD-1/PD-L1 isPLA positivity. Analyses are shown for intra-tumoral, peri-tumoral and combined regions.

## References

1. Aliazis K, Christofides A, Shah R, Yeo YY, Jiang S, Charest A, et al. The tumor microenvironment’s role in the response to immune checkpoint blockade. Nat Cancer. 2025 Jun;6(6):924–37. doi:10.1038/s43018-025-00986-3 PubMed PMID: 40514448; PubMed Central PMCID: PMC12317369.

2. Maiorano BA, Di Maio M, Cerbone L, Maiello E, Procopio G, Roviello G, et al. Significance of PD-L1 in Metastatic Urothelial Carcinoma Treated With Immune Checkpoint Inhibitors: A Systematic Review and Meta-Analysis. JAMA Netw Open. 2024 Mar 4;7(3):e241215. doi:10.1001/jamanetworkopen.2024.1215 PubMed PMID: 38446479; PubMed Central PMCID: PMC10918499.

3. Balar AV, Galsky MD, Rosenberg JE, Powles T, Petrylak DP, Bellmunt J, et al. Atezolizumab as first-line treatment in cisplatin-ineligible patients with locally advanced and metastatic urothelial carcinoma: a single-arm, multicentre, phase 2 trial. Lancet Lond Engl. 2017 Jan 7;389(10064):67–76. doi:10.1016/S0140-6736(16)32455-2 PubMed PMID: 27939400; PubMed Central PMCID: PMC5568632.

4. Vuky J, Balar AV, Castellano D, O’Donnell PH, Grivas P, Bellmunt J, et al. Long-Term Outcomes in KEYNOTE-052: Phase II Study Investigating First-Line Pembrolizumab in Cisplatin-Ineligible Patients With Locally Advanced or Metastatic Urothelial Cancer. J Clin Oncol Off J Am Soc Clin Oncol. 2020 Aug 10;38(23):2658–66. doi:10.1200/JCO.19.01213 PubMed PMID: 32552471.

5. Bellmunt J, de Wit R, Vaughn DJ, Fradet Y, Lee JL, Fong L, et al. Pembrolizumab as Second-Line Therapy for Advanced Urothelial Carcinoma. N Engl J Med. 2017 Mar 16;376(11):1015–26. doi:10.1056/NEJMoa1613683 PubMed PMID: 28212060; PubMed Central PMCID: PMC5635424.

6. Lindberg A, Muhl L, Yu H, Hellberg L, Artursson R, Friedrich J, et al. In Situ Detection of Programmed Cell Death Protein 1 and Programmed Death Ligand 1 Interactions as a Functional Predictor for Response to Immune Checkpoint Inhibition in NSCLC. J Thorac Oncol Off Publ Int Assoc Study Lung Cancer. 2025 May;20(5):625–40. doi:10.1016/j.jtho.2024.12.026

7. Sánchez-Magraner L, Gumuzio J, Miles J, Quimi N, Martínez Del Prado P, Abad-Villar MT, et al. Functional Engagement of the PD-1/PD-L1 Complex But Not PD-L1 Expression Is Highly Predictive of Patient Response to Immunotherapy in Non–Small-Cell Lung Cancer. J Clin Oncol. 2023 May 10;41(14):2561–70. doi:10.1200/JCO.22.01748

8. Powles T, Walker J, Andrew Williams J, Bellmunt J. The evolving role of PD-L1 testing in patients with metastatic urothelial carcinoma. Cancer Treat Rev. 2020 Jan;82:101925. doi:10.1016/j.ctrv.2019.101925 PubMed PMID: 31785413.

9. Mariathasan S, Turley SJ, Nickles D, Castiglioni A, Yuen K, Wang Y, et al. TGFβ attenuates tumour response to PD-L1 blockade by contributing to exclusion of T cells. Nature. 2018 Feb;554(7693):544–8. doi:10.1038/nature25501

10. Zhao C, Germain RN. Multiplex imaging in immuno-oncology. J Immunother Cancer. 2023 Oct;11(10):e006923. doi:10.1136/jitc-2023-006923

11. Lin JR, Izar B, Wang S, Yapp C, Mei S, Shah PM, et al. Highly multiplexed immunofluorescence imaging of human tissues and tumors using t-CyCIF and conventional optical microscopes. eLife. 2018 Jul 11;7:e31657. doi:10.7554/eLife.31657

12. Bollhagen A, Bodenmiller B. Highly Multiplexed Tissue Imaging in Precision Oncology and Translational Cancer Research. Cancer Discov. 2024 Nov 1;14(11):2071–88. doi:10.1158/2159-8290.CD-23-1165

13. Black S, Phillips D, Hickey JW, Kennedy-Darling J, Venkataraaman VG, Samusik N, et al. CODEX multiplexed tissue imaging with DNA-conjugated antibodies. Nat Protoc. 2021 Aug;16(8):3802–35. doi:10.1038/s41596-021-00556-8

14. Method of the Year 2024: spatial proteomics. Nat Methods. 2024 Dec;21(12):2195–6. doi:10.1038/s41592-024-02565-3

15. Söderberg O, Gullberg M, Jarvius M, Ridderstråle K, Leuchowius KJ, Jarvius J, et al. Direct observation of individual endogenous protein complexes in situ by proximity ligation. Nat Methods. 2006 Dec;3(12):995–1000. doi:10.1038/nmeth947

16. Klaesson A, Grannas K, Ebai T, Heldin J, Koos B, Leino M, et al. Improved efficiency of in situ protein analysis by proximity ligation using UnFold probes. Sci Rep. 2018 Mar 29;8(1):5400. doi:10.1038/s41598-018-23582-1 PubMed PMID: 29599435; PubMed Central PMCID: PMC5876389.

17. Mardamshina M, Ballllosera Navarro F, Martinez Casals A, Avenel C, Wählby C, Lundberg E. Streamlining Multiplexed Tissue Image Analysis with PIPΣX: An Integrated Automated Pipeline for Image Processing and EXploration for Diverse Tissue Types [Internet]. 2025 [cited 2025 Nov 3]. Available from: http://biorxiv.org/lookup/doi/10.1101/2025.05.04.652145 doi:10.1101/2025.05.04.652145

18. Pielawski N, Andersson A, Avenel C, Behanova A, Chelebian E, Klemm A, et al. TissUUmaps 3: Improvements in interactive visualization, exploration, and quality assessment of large-scale spatial omics data. Heliyon. 2023 May;9(5):e15306. doi:10.1016/j.heliyon.2023.e15306 PubMed PMID: 37131430; PubMed Central PMCID: PMC10149187.

19. Solorzano L, Partel G, Wählby C. TissUUmaps: interactive visualization of large-scale spatial gene expression and tissue morphology data. Mathelier A, editor. Bioinformatics. 2020 Aug 1;36(15):4363–5. doi:10.1093/bioinformatics/btaa541

20. Oliveira G, Stromhaug K, Klaeger S, Kula T, Frederick DT, Le PM, et al. Phenotype, specificity and avidity of antitumour CD8+ T cells in melanoma. Nature. 2021 Aug;596(7870):119–25. doi:10.1038/s41586-021-03704-y PubMed PMID: 34290406; PubMed Central PMCID: PMC9187974.

21. Aung TN, Monkman J, Warrell J, Vathiotis I, Bates KM, Gavrielatou N, et al. Spatial signatures for predicting immunotherapy outcomes using multi-omics in non-small cell lung cancer. Nat Genet. 2025 Oct;57(10):2482–93. doi:10.1038/s41588-025-02351-7

22. Yamaguchi H, Hsu JM, Sun L, Wang SC, Hung MC. Advances and prospects of biomarkers for immune checkpoint inhibitors. Cell Rep Med. 2024 Jul 16;5(7):101621. doi:10.1016/j.xcrm.2024.101621 PubMed PMID: 38906149; PubMed Central PMCID: PMC11293349.

23. Hjorthén G, Costa Svedman F, Holmsten K, Ullén A. Impact of glucocorticoid treatment and clinical prognostic factors for outcome in patients with advanced urothelial cancer treated with pembrolizumab. Urol Oncol. 2025 Aug;43(8):467.e21–467.e29. doi:10.1016/j.urolonc.2025.02.017 PubMed PMID: 40140265.

24. Deen AJ, Thorsson J, O’Roberts EM, Panshikar P, Ullman T, Krantz D, et al. Making Multiplexed Imaging Flexible: Combining Essential Markers With Established Antibody Panels. J Histochem Cytochem Off J Histochem Soc. 2024;72(8–9):517–44. doi:10.1369/00221554241274856 PubMed PMID: 39215640; PubMed Central PMCID: PMC11421402.

25. PhenoCycler-Fusion User Guide 2.2.0 [Internet]. Available from: https://www.akoyabio.com/phenoimager-fusion-software-2-2-0/

26. NaveniFlex Bond RX Atto647N [Internet]. Available from: https://navinci.se/wp-content/uploads/2025/06/IFU-0026-NaveniFlex-BOND.pdf

